# RANKL inhibition with denosumab improves fibrous dysplasia by decreasing lesional cell proliferation and increasing osteogenesis

**DOI:** 10.1101/2022.10.24.22281375

**Authors:** Luis F de Castro, Jarred M Whitlock, Zachary Michel, Kristen Pan, Jocelyn Taylor, Vivian Szymczuk, Sriram S Paravastu, Babak Saboury, Georgios Z Papadakis, Leonid V Chernomordik, Xiaobai Li, Kelly Milligan, Brendan Boyce, Scott Paul, Daniel Martin, Michael T Collins, Alison M Boyce

## Abstract

**BACKGROUND:** Fibrous dysplasia (FD) is a rare, disabling disease with no established treatments. Growing evidence supports inhibiting the pro-osteoclastic factor receptor activator of nuclear Kappa-B ligand (RANKL) as a potential treatment strategy. We conducted a phase 2 trial evaluating the anti-RANKL drug denosumab in adults with FD, with an emphasis on investigating post-discontinuation bone turnover rebound, and cellular mechanisms underlying anti-RANKL effects on FD osteoprogenitors.

**METHODS:** Eight subjects received denosumab for 6-months and were observed for 8-months post-discontinuation. Efficacy and safety were evaluated using bone turnover markers, ^18^F-NaF PET/CT, and lesion biopsies. RANKL neutralization effects were assessed by histology, RNASeq, and an FD mouse model. Interplay between osteoclasts and FD osteoprogenitors was assessed in an *ex vivo* lesion model.

**RESULTS:** Denosumab markedly reduced bone turnover and radiographic lesional activity in all subjects. Denosumab was well-tolerated during the treatment period, however post-discontinuation turnover reached or exceeded pre-treatment in six subjects, associated with severe hypercalcemia in one. Histology and whole-exome RNA sequencing showed reduced FD cell proliferation and increased osteogenic maturation, with increased lesional bone formation. The *ex vivo* model supported the dependence of FD cell proliferation on osteoclast activation.

**CONCLUSIONS:** Osteoclast inhibition by anti-RANKL decreased FD cell proliferation and lesional activity, enabling osteogenic maturation and bone formation. These findings provide new understanding of FD pathogenesis as driven by crosstalk between osteoclasts and pre-osteoblast/osteoblasts, and support denosumab as a mechanistically-driven treatment strategy. Marked bone turnover rebound with post-discontinuation hypercalcemia occurs in a subset of patients, particularly younger individuals with high disease burden.

**TRIAL REGISTRATION:** ClinicalTrials.gov NCT03571191

**FUNDING:** This work was supported by the Intramural Research Program of the NIDCR, NICHD, and Clinical Center, National Institutes of Health. Clinical trial NCT03571191 was conducted as an investigator-sponsored study supported by Amgen, Inc. This research was supported in part by the NIDCR Genomics and Computational Biology Core: ZIC DC000086 and Veterinary Resources Core: ZIC DE000740-05. Work in MTC lab and LVC labs were supported by the of Research on Women’s Health (ORWH) through the Bench to Bedside Program award #884515.

## INTRODUCTION

Fibrous dysplasia (FD) is a rare, mosaic disorder leading to fractures, pain, and disability. Gain-of-function *GNAS* variants alter skeletal stem cell differentiation, leading to expansile lesions of highly proliferative, partially mineralized fibro-osseus tissue ^1^. Disease presents along a broad spectrum, and may occur in association with extraskeletal features, termed McCune-Albright syndrome ^2^. In the craniofacial skeleton, FD lesion expansion causes facial asymmetry and functional deficits, such as vision and hearing loss. Lesions in weight-bearing bones can fracture and deform, leading to pain and ambulation impairment. There are no approved treatments, and there is a critical need for targeted therapies.

The development of effective treatments in FD has been hindered by knowledge gaps in the interplay between lesional cell populations. Abnormal osteogenic cells comprise the dominant lineage and lead to the signature histopathologic features of FD, including fibrosis, altered bone formation, and loss of marrow structures ^1^. Osteoclasts are variably prominent in FD tissue, however their role in establishing and maintaining FD lesions remains unknown. Bisphosphonates, a class of antiresorptive drugs that attach to hydroxyapatite and impair osteoclast activity, have been studied for decades in FD, but show no convincing effects on lesional activity or microarchitecture ^3-5^. Growing evidence suggests that targeting receptor activator of nuclear Kappa-B ligand (RANKL), a cytokine that promotes osteoclastogenesis, may be a more promising strategy. In current understanding of physiologic bone remodeling, RANKL release by osteoprogenitors and other cell types binds RANK on the surface of osteoclast precursors, promoting fusion into mature osteoclasts. RANKL is highly expressed in FD cells, and induces osteoclastogenesis in co-cultured monocytes ^6,7^. Murine models also show that RANKL inhibition arrests expansion and improves mineralization of skeletal lesions ^8,9^.

Denosumab is a monoclonal antibody to RANKL approved for treatment of osteoporosis and bone tumors ^10,11^, and retrospective studies suggest it has favorable effects on FD expansion and activity ^6,12,13^. However, important questions remain about efficacy and safety. The effects of denosumab on FD lesional composition in humans have not been evaluated, and effects of osteoclast inhibition on FD cells have not been established. The antiresorptive action of denosumab is transient, and drug discontinuation has been associated with rebound increased bone turnover, with hypercalcemia reported in 2 FD patients ^6,13^. There is thus a critical need to define the lesional effects, duration of activity, and safety of denosumab treatment and discontinuation.

Here we report the results of a phase 2 study of denosumab in adults with FD. We assessed effects of RANKL inhibition in FD tissue and in pre-clinical models to explore cellular effects on FD osteoprogenitors.

## METHODS

### Trial design and oversight

A phase 2 open-label study was conducted at the NIH (NCT03571191). This investigator sponsored study was supported by Amgen, Inc; study design, conduct, and analyses were performed by the investigators. The trial was approved by the NIH Investigational Review Board, and informed consent was obtained from all subjects. The study was monitored by a data safety and monitoring committee organized by the National Institute of Dental and Craniofacial Research.

Subjects received denosumab for 6-months at a dose of 120 mg every 4 weeks, with loading doses on weeks 2 and 3 ^11^. Subjects were monitored for 8-months post-denosumab discontinuation. Study procedures were conducted from June 2019 to July 2022. The primary objective was to determine the effect of denosumab on bone turnover. Secondary objectives included effects on lesional activity, pain, discontinuation effects on bone turnover rebound, and safety. Eligibility criteria are included in Table S1.

### Clinical endpoint collection

^18^F-NaF PET/CT scans were performed using Siemens Biograph 128_mCT models (Siemens Medical Solutions USA, Malvern, USA) and read by 2 central readers (BS, GZP). Standard correlation analysis between reads, and agreement between readers using Bland-Altman analysis, was performed. Minimally-invasive percutaneous FD lesion biopsies were performed at baseline and 6-months (Table 1). Tissues were evaluated for histology, protein, and mRNA expression (Supplementary appendix).

**Table 1.** Subject Demographics

| Subject ID | Age (years) | Sex | Skeletal Burden Score<sup>1</sup> | Regions of FD involvement | MAS endocrinopathies | Biopsy locations |
| --- | --- | --- | --- | --- | --- | --- |
| DB01 | 18-25 | female | 7 | Craniofacial | None | Maxilla<sup>2</sup> |
| DB02 | 51-55 | female | 39 | Craniofacial, axial, appendicular | Precocious puberty | Rib |
| DB03 | 26-30 | female | 72 | Craniofacial, axial, appendicular | Precocious puberty | Iliac crest<sup>3</sup> |
| DB04 | 26-30 | female | 17 | Craniofacial, axial | GH excess | Rib |
| DB05 | 31-35 | female | 27 | Craniofacial, axial, appendicular | Precocious puberty | Rib |
| DB06 | 18-25 | female | 64 | Craniofacial, axial, appendicular | Precocious puberty, hyperthyroidism, GH excess | Rib<sup>4</sup> |
| DB07 | 36-40 | female | 30 | Craniofacial, axial, appendicular | Precocious puberty | Scapula |
| DB08 | 23-30 | female | 69 | Craniofacial, axial, appendicular | Precocious puberty | Iliac crest |
| DB09 | 18-25 | female | 36 | Craniofacial, axial, appendicular | Precocious puberty | Rib |

<sup>1</sup>Validated semi-quantitative measure defining the proportion of the skeleton involved with FD. Values range from 0 (no FD) to 75 (panostotic FD). See Collins et al, J Bone Miner Res 2005 Feb;20(2):219-26.
<sup>2</sup>Post-treatment biopsy deferred due to potential risk of inducing osteonecrosis of the jaw. <sup>3</sup>Post-treatment biopsy deferred due to intercurrent illness, unrelated to study intervention. <sup>4</sup>Post-treatment biopsy deferred after subject was withdrawn from the study.

### *In vivo* and *ex vivo* models of Fibrous Dysplasia

Mice expressing the Gα _s_^R201C^ variant were generated as described ^14^. 10 week old mice were induced with doxycycline, and treated with anti-RANKL antibody or non-immune isotype per the Supplementary appendix. Plasma was collected weekly, and on day 58 mice were euthanized and tissue was collected. Experiments were conducted under a protocol approved by the NIH/NIDCR Animal Care and Use committee.

Gα _s_^R201C^ expression was induced in *ex vivo* bone marrow cultures ^15^ and then treated with anti-RANKL antibody. MC3T3-E1 clone 4 and 14 cells were cultured as previously described ^16^. Additional methods are included in the Supplementary appendix.

### Statistical analyses

For clinical outcome variables, changes from baseline to 6-months were assessed with one-sample t-tests or Wilcoxon signed-rank tests. Medians (IQR, range) are reported unless otherwise indicated. Bone turnover marker analyses are reported as both absolute and percent change from baseline. Data analyses were conducted in SAS (the SAS Institute, Cary, NC) by XL. *In vivo* mouse analyses were performed using non-paired t-tests or Mann-Whitney tests. For longitudinal measures, paired t-tests compared baseline to timepoint values. Mouse data are reported as average and standard deviation when shown, unless otherwise indicated, and analyses were conducted on GraphPad Prism

8.0.2 by LFdC. Culture experiments were evaluated by t-tests paired by donor mouse, and non-paired t-tests for MC3T3-E1 experiments by JW.

## RESULTS

### Denosumab as a therapeutic intervention in FD: treatment period

Nine adults were enrolled and received denosumab (Table 1, Fig S1). One subject (DB06) was withdrawn following the first dose after experiencing prolonged hypocalcemia with inappropriately normal parathyroid hormone levels, consistent with previously unrecognized partial post-surgical hypoparathyroidism. Eight subjects completed the 6-month treatment period, and denosumab was otherwise well-tolerated. All subjects received supplemental calcium and calcitriol. Six developed mild-moderate asymptomatic hypocalcemia, managed with oral supplementation and deemed related to denosumab. There were no other adverse events during the treatment period that were related to denosumab.

Treatment was associated with marked declines in the bone formation marker P1NP (−92%, range -87 to -93%, p<0.001), and the resorption marker CTX (−82%, range -61 to -86%, p<0.001)(Fig 1A&B). ^18^F-NaF PET/CT scans likewise showed marked reduction in lesional activity measures Global Burden of Disease and Total Lesion Volume (Fig 1C-E).

**Figure 1.**
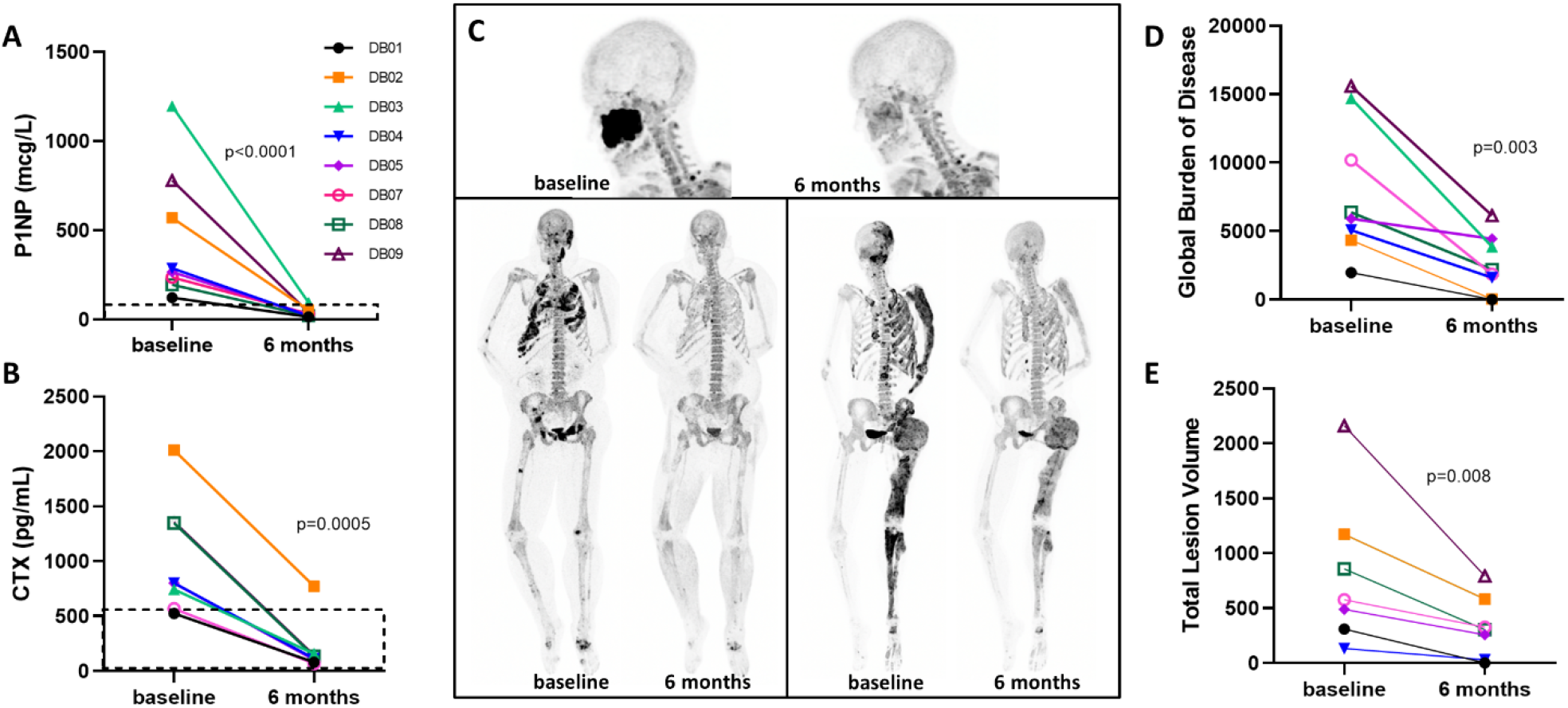
Effect of denosumab on bone turnover markers and radiographic lesion activity. The left-hand panels show individual changes in the bone formation marker P1NP (A) and the bone resorption marker CTX (B). The dotted lines indicate the normal range for each assay. Panel C shows representative images of ^18^F-NaF PET/CT scans, depicting qualitative changes in FD lesion tracer uptake in subjects DB01 (top), DB02 (left), and DB09 (right). The right-hand panels show individual changes in quantitative measures of lesion activity, including Global Burden of Disease, determined as the product of SUVmean multiplied by total volume of all ^18^F-NaF avid FD lesions, and Total Lesion Volume, defined as summation of the volumes of all ^18^F-NaF avid FD lesions.

Denosumab was associated with reduction in worst pain score [6.4 (IQR 3.25) to 3.5 (IQR 6.25), p=0.02] and pain interference [4.5 (IQR 8.25) to 3 (IQR 9.75), p=0.03], as assessed by the Brief Pain Inventory. The 9-minute walk test showed no change (assessed in 6 ambulatory patients). Two subjects had objective improvement in severe FD-related morbidity. DB02 had expansile rib FD (Fig 1C) with severely restrictive lung disease, necessitating continuous supplemental oxygen. After starting denosumab pulmonary function improved and oxygen was discontinued (Table S2). DB04 had aggressive craniofacial FD resulting in recurrent visual impairment, necessitating 8 optic canal decompression surgeries in the preceding 4 years. After starting denosumab, her ophthalmologic exam stabilized and vision has been preserved.

### Denosumab as a therapeutic intervention in FD: discontinuation period

Eight subjects were monitored for 8-months post-denosumab discontinuation. All subjects received zoledronic acid 4 weeks following the final denosumab dose, with subsequent infusions as clinically-indicated (Table S3). CTX rebounded above pre-treatment levels in 4 subjects, 3 to 8 months post-discontinuation (Fig 2, Table S3). Hypercalcemia developed in 3 subjects. In one (DB03) hypercalcemia was severe, peaking at 23 mg/dL (normal 8.6-10.2) 3-months post-discontinuation. The subject was treated with inpatient intravenous fluids, calcitonin, zoledronic acid, and an additional 60 mg of denosumab, and recovered without sequelae. The additional 2 cases of hypercalcemia were mild and asymptomatic.

**Figure 2.**
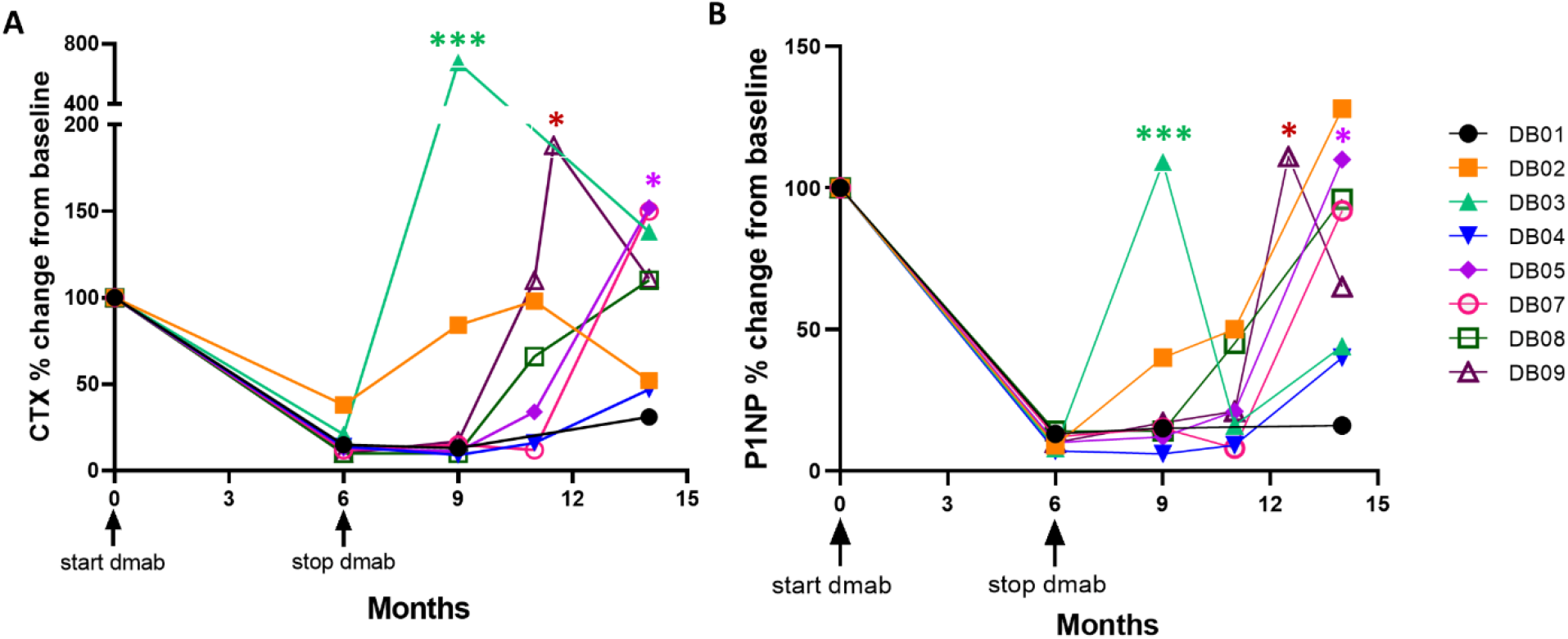
Effects of denosumab discontinuation on bone turnover markers. Graphs show individual percent changes from baseline in the resorption marker CTX (A) and formation marker P1NP (B). *** corresponds to severe hypercalcemia, * corresponds to mild-moderate hypercalcemia.

### Denosumab decreased cellularity and increased osteogenic maturation of FD tissue

As expected, TRAP+ or RANK+ osteoclasts were absent in FD tissue after denosumab treatment, although RANK+ mononuclear precursors remained (Figs 3A&B, S2). Treatment caused a marked increase in lesion mineralization and a decrease in FD cellularity, as measured in H&E stains (Fig 3A&B, S3). Reduced cellular positivity for MCM2, and increased SOST positive osteocytes confirmed that osteoclastogenesis blockade led to reduced proliferation and increased maturation of FD cells. Earlier osteoblastic differentiation markers RUNX2 and OCN did not change significantly post-denosumab and mRNA hybridization analysis of Gα _s_^R201C^ indicated a decrease in lesional mutation expression (Figs 3A&B, S2, S4).

**Figure 3.**
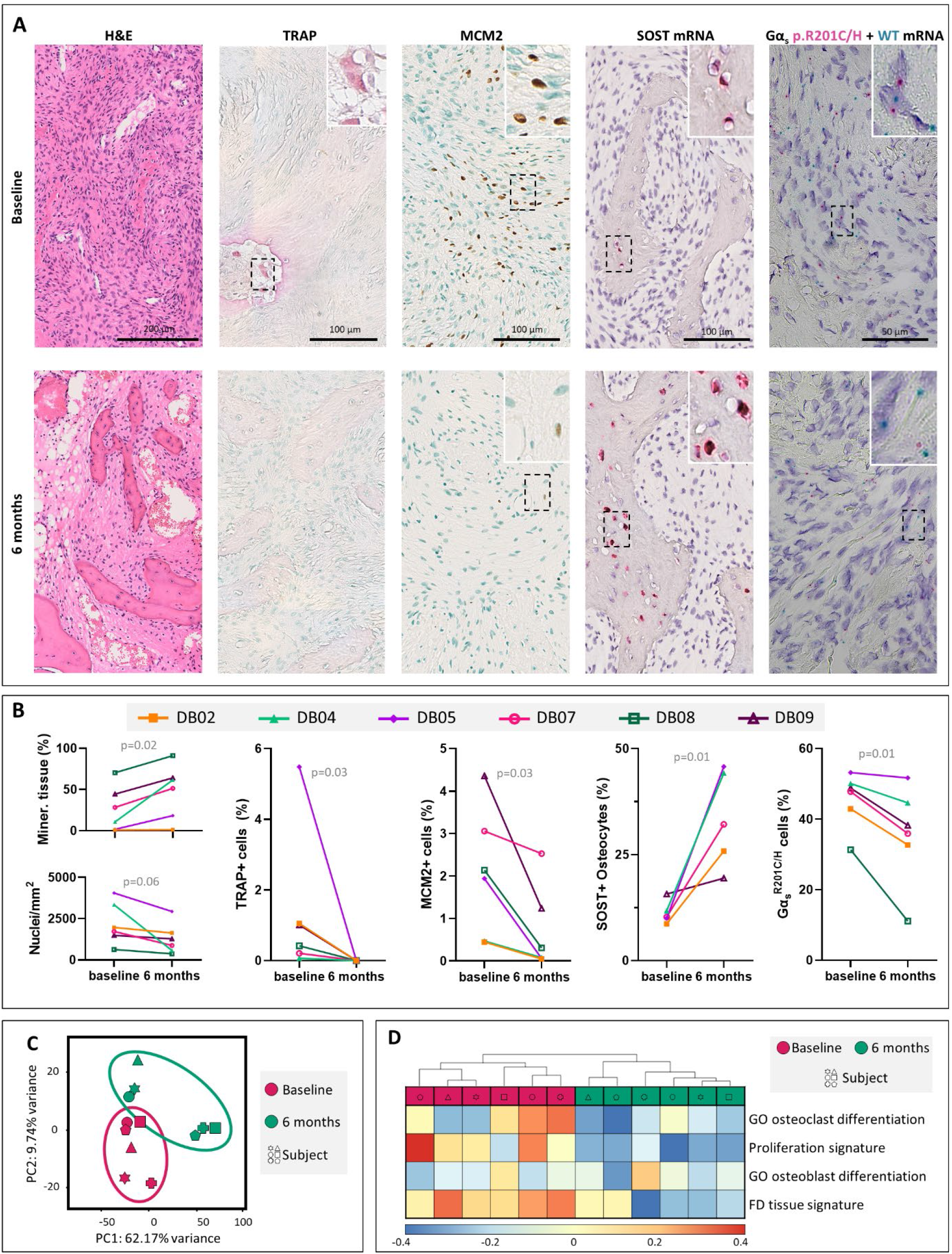
Histological and molecular changes in human FD post-denosumab treatment. A) Representative images of changes in FD tissue histology before (baseline) and after treatment with denosumab (6 months). H&E from subject DB04 showing increased lesional mineralization and decreased cellularity. Absence of Trap activity (DB07). Decreased proliferation by Mcm2 proliferation marker (DB02). Upregulation in sclerostin mRNA expression (DB07). I) Decreased mutation burden, labelling individual molecules of WT Gα_s_ mRNA (green dots) and Gα_s_^R201C^ mRNA (red dots) in subject DB04. B) Quantification of the stains in A. H&E nuclear density and bone tissue area, TRAP+ cells showing no post-treatment osteoclasts, decreased MCM2+ cells, increased SOST+ osteocytes and decreased relative amount of Gα _s_^R201C^ -expressing cells. C) RNAseq principal component (PC) analysis showing segregation of the transcriptomic profiles of human FD tissue before and after denosumab treatment. D) Unsupervised clustering of whole transcriptome GSVA scores against selected molecular signatures and genesets.

Transcriptomic principal component analysis showed segregation of the biopsies according to treatment status rather than donor identity, and confirmed the changes in osteoclastogenesis, proliferation, and osteoblast differentiation observed in histology (Fig 3C&D, S5). The transcriptome of mouse ulna FD tissue was compared to wildtype bone to develop a FD tissue genetic signature; the transcriptome of denosumab-treated human FD in comparison to pre-treated tissue showed an inverse regulation of this signature (Fig 3D, Table S4).

### *In vivo* and *ex vivo* FD models supported and expanded the observations in human biopsies

A doxycycline-inducible mouse model of FD was used to interrogate lesional and cellular effects of anti-RANKL treatment. Mice developed mild FD lesions in their appendicular skeleton for 4 weeks, received αRANKL or isotype control antibodies for 4 additional weeks, and were euthanized two days after the last injection. Disease progression was measured longitudinally using a quantitative X-ray score and plasma bone turnover markers TRAP5b and P1NP (Figs 4A, S7, Table S5). Trap5b and P1NP increased above baseline levels one week after disease induction and dropped after αRANKL treatment, reaching lower than physiological levels on weeks one and three post-treatment start, respectively. µCT analysis of FD lesions showed decreased mineral density post-induction, with abundant partially mineralized abnormal bone. αRANKL treatment reverted these changes, showing higher than normal volume of mineralized tissue (Figs 4B, S6).

**Figure 4.**
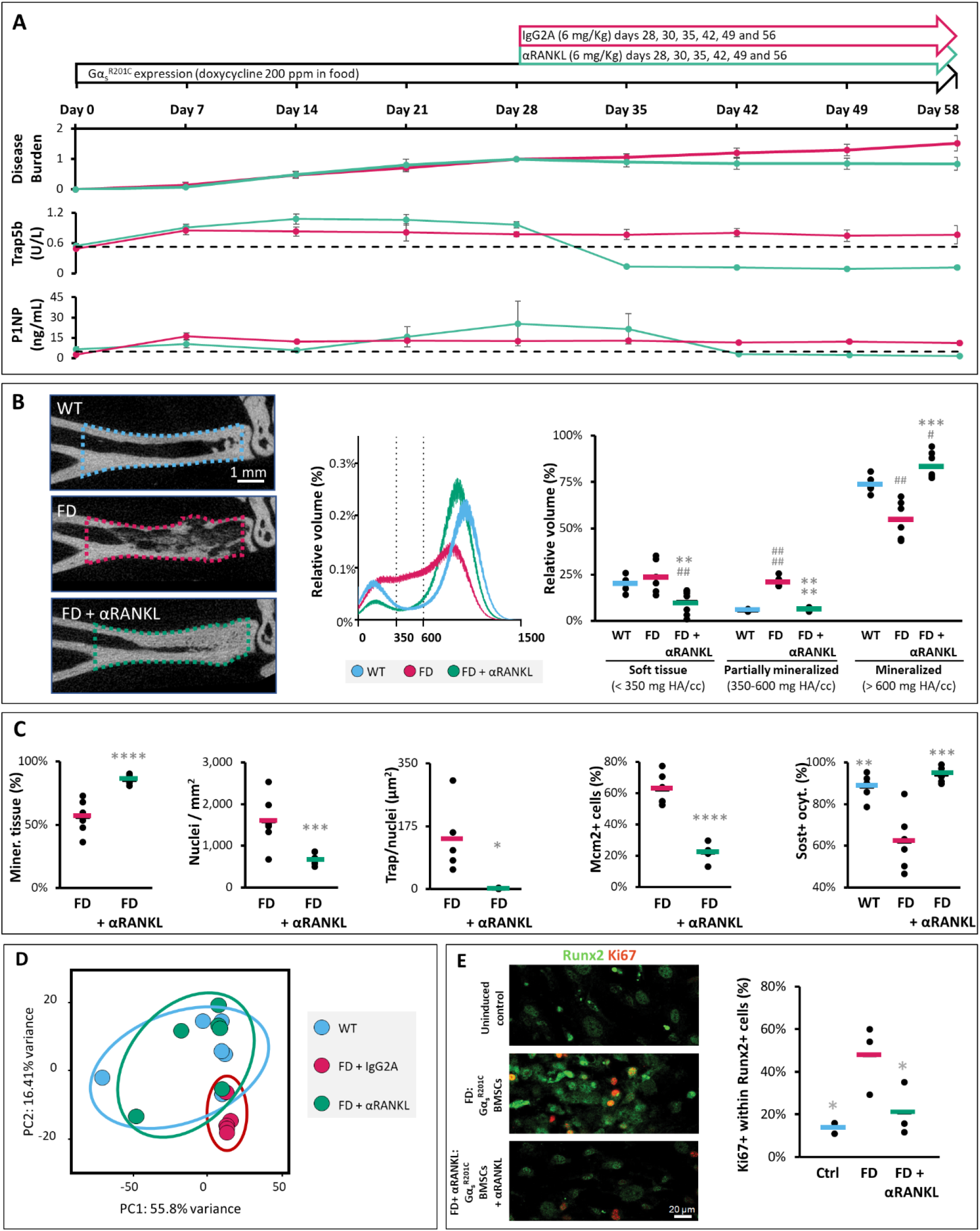
αRANKL-mediated inhibition of osteoclastogenesis arrests the proliferation of FD osteoprogenitors *in vivo* and *ex vivo*, enabling their maturation and tissue ossification. A) Longitudinal FD mice disease burden survey by lesional X-ray scores. Data was normalized against each lesional score at the start of αRANKL/isotype control treatment, 28 days after FD lesion induction by doxycycline administration to the mice. Serum Trap5b and P1NP are markers of bone resorption and formation respectively. The black dotted line is the average baseline value. B) uCT scan slice showing the lesional volume of interest analyzed projected in a mid-sagittal section of the distal tibia. Distribution histogram of the density values in the volume of interest, showing group averages. FD tissue is characterized by the appearance of a shoulder in values that correspond to partial mineralization. 350 and 600 mg HA/cc density values were used to threshold soft, partially mineralized and mineralized tissue. The panel to the left shows the relative volumes of the three tissue categories in each mouse (black dots) and group averages (bars). C) Histological and immunohistological evaluation of mouse FD and control tissue replicating the observations in human FD after neutralizing RANKL. D) Principal component (PC) analysis of mouse FD tissue, showing segregation of the αRANKL-treated FD tissue group and overlapping with site-matched WT tissue. E) *ex vivo* FD microenvironment model. Normal bone marrow is plated and bone marrow stromal cells (BMSCs, osteoprogenitors) are induced to express Gα _s_^R201C^ by doxycycline administration, which leads to the formation of osteoclasts from wildtype precursors that, in turn, promote FD BMSCs proliferation. When αRANKL is administered to the culture, there is a full inhibition of BMSCs proliferation, as shown by the decreased positivity for Ki67 of Runx2+ BMSCs. In (A) X-ray scores are significantly different from day 35, all Trap5b measurements were significantly different to baseline at all time points and different between groups from day 35. P1NP is significantly different than baseline values in the untreated group on days 7 and 49 and in the treated group on days 49 and 56. Treated and untreated groups are significantly different from day 42. * is p<0.05, ** is p<0.01, *** is p<0.001 and **** is p<0.0001 vs FD group. # is p<0.05, ## is p<0.01 and #### is p<0.0001 vs control group.

All the histological changes observed in human FD biopsies post-denosumab treatment were confirmed in mouse FD tissue. Mouse FD bone demonstrated lower than normal osteocyte Sost positivity. Similar to human FD, αRANKL treatment normalized Sost expression, but also decreased Runx2 and ALP staining, markers of committed osteoprogenitors and osteoblasts, in mouse FD (Figs 4C, S8, S9). Transcriptomic principal component analysis showed overlap between αRANKL-treated FD and unaffected bone, and supported all findings observed in human biopsies except increased proliferation in FD tissue and its reversion by αRANKL (Figs 4D, S10, table S4).

An *ex vivo* model of FD was developed, in which mouse bone marrow containing bone marrow stromal cells (BMSCs, osteoprogenitors) and monocyte/macrophage osteoclast progenitors was cultured and induced with doxycycline for Gα _s_^R201C^ expression by only BMSCs. This model suggested strict dependance of osteoprogenitors on osteoclastogenesis to support cell proliferation. After induction of BMSCs with doxycycline, functional wildtype osteoclasts appear in these cultures by fusion of mononuclear precursors, and release Rank containing extracellular vesicles, while Runx2+ BMSCs increase their positivity for the Ki67 proliferation marker (Fig 4E, S11A-F). When αRANKL is added, this process is inhibited and BMSCs stop proliferating, despite a lack of direct effect on Ki67 positivity in wildtype MC3T3-E1 osteoprogenitors (Fig S11G).

## DISCUSSION

In this study, denosumab treatment in patients with FD had potent inhibitory effects on bone turnover and lesional activity, mediated by changes in proliferation and osteogenic differentiation of FD cells. Denosumab thus has marked effects on FD lesion composition beyond its expected antiresorptive activity. Taken together, these findings establish the critical importance of crosstalk between osteoclastic and osteogenic cells in FD pathogenesis, and support targeting RANKL as an effective therapeutic approach.

These alterations in FD lesional composition represent clinically important changes, because a) FD cell proliferation is associated with increased lesional activity and expansion ^4,7,17^, and b) reduced mineralization is associated with bone weakness, deformities, and fractures ^18,19^. While RANK/RANKL signaling in bone remodeling has been characterized as pro-osteoclastogenic, recent work by Ikebuchi et al suggests RANKL may actually have a bidirectional role, reporting promotion of osteoblast differentiation by osteoclast-released RANK+ vesicles through interaction with membrane-anchored RANKL ^20^. In our study, osteoclast formation in FD *ex vivo* was accompanied by release of extracellular vesicles and increased FD cell proliferation, changes that were fully inhibited with anti-RANKL treatment. These findings thus lend support to this new paradigm for osteoclast-osteoblast coupling, and point to this cross-talk as an essential pathogenic mechanism in FD that is successfully targeted by RANKL inhibition. Further studies, including characterization of the content and production of osteoclast-derived vesicles, are needed to confirm and expand these findings.

Denosumab had potent effects on radiographic markers of FD lesional activity in all subjects. ^18^F-NaF is a bone-seeking radiopharmaceutical in which fluoride exchanges with hydroxyl on hydroxyapatite surfaces, and previous work demonstrates significant correlations with clinically meaningful outcomes, including fractures, surgeries, deformities, and functional impairment ^21^. Taken together with the histopathologic analyses, these findings indicate that ^18^F-NaF PET/CT is an effective surrogate marker to reflect therapeutic changes in FD tissue.

Histopathologic and ^18^F-NaF findings were accompanied by similarly marked changes in serum bone turnover markers, which declined largely into the normal range. Interestingly, despite its established anti-osteoclastic effects, denosumab had a more potent effect on P1NP (a bone formation marker) than CTX (a bone resorption marker). This is in contrast to a controlled trial of the bisphosphonate alendronate in FD, which demonstrated effects on resorption markers alone ^3^. Inefficacy of bisphosphonates in FD may result because their action requires incorporation into mineralizing matrix, which is significantly diminished in FD. It is unknown if the increased lesional mineral formation generated by denosumab may subsequently enable incorporation of bisphosphonates into FD lesions. If so, it could potentially open up a therapeutic role for bisphosphonates, which (unlike denosumab) have the advantage of sustained anti-resorptive effects. Further studies are needed to investigate the feasibility and safety of potential co-treatment regimens.

As expected, effects of denosumab were transient and reversible after treatment discontinuation. The duration of efficacy was variable, and by 8-months post-discontinuation, turnover markers reached or exceeded pre-treatment levels in 6 of 8 subjects. The onset of rebound ranged from 3 to 8 months post-discontinuation, and was associated with severe hypercalcemia in 1 subject, highlighting an important and potentially life-threatening complication. Previous reports of post-discontinuation hypercalcemia in FD have been limited to 2 children ^6,13^, and a retrospective review of clinically-treated adults did not identify symptomatic hypercalcemia ^22^. In our study, the subject with severe hypercalcemia had extremely elevated baseline bone turnover, with the highest Skeletal Burden Score and P1NP level in the cohort. Similarly, 2 subjects with the lowest Skeletal Burden Scores maintained bone turnover markers below pre-treatment levels 8-months post-discontinuation. Taken together, these findings indicate clinicians should be judicious in utilizing denosumab for FD, and should proceed with particular caution in younger patients and those with high disease burden. These findings also suggest that optimal use of denosumab likely necessitates an individualized approach, because patients with higher baseline bone turnover require more frequent dosing to maintain bone turnover suppression.

The results of this study are promising, particularly given the marked response in all subjects with short-term treatment. However this study is small, and findings need to be extended in larger and more diverse cohorts. Additional studies are also needed to define optimal dosing regimens and the safety of long-term treatment in FD. This is particularly important in childhood, which is the period during which FD lesions expand and disease burden is established ^17^. A trial of denosumab to prevent FD lesion progression in children is ongoing (NCT05419050); based on findings reported here, this trial will use frequent low-dose intervals in anticipation of the likely need for long-term treatment.

In conclusion, this study demonstrates that beyond its expected anti-osteoclastic effects, denosumab reduces FD lesion activity by decreasing FD cell proliferation and increasing osteogenic maturation, leading to increased lesional bone formation. These findings provide new understanding of FD pathogenesis as driven by crosstalk between osteoclasts and pre-osteoblast/osteoblasts, and support denosumab as a mechanistically driven treatment strategy. Vigorous bone turnover rebound with the potential for post-discontinuation hypercalcemia occurs in a subset of patients, and further study is needed to define optimal dosing strategies, particularly in younger individuals with high skeletal disease burden.

## Data Availability

All data produced in the present study are available upon reasonable request to the authors

## AUTHOR CONTRIBUTIONS

Patient recruitment, clinical information collection, specimen collection, and study coordination were done by AB, KM, MC, and SP. Experimental design and interpretation were done by LFdC, MC, LC, JW, and AB. Histological examination and quantification was performed by LFdC, KP, ZM, JT, VS, and BB. Ex vivo culture model and experiments employing it were developed and designed by JW and LC. Experiments were carried out and analyzed by JW. Bone specimen processing, RNA extraction, and bioinformatic analyses were done by LFdC, AB, ZM, and DM. Imaging analyses were done by GP, SP, and BS. Statistical analyses were performed by XL. The manuscript was written by LFdC and AB with input from all authors.

## ACKNOWLEDGEMENTS

We are grateful to individuals with FD and their families who participated in this study. We thank the NIH endocrinology fellows for providing clinical care. We are grateful to Tiffani Slaughter, Rebeca Galisteo, and the Veterinary Research Core at NIDCR for their continuous support in the animal experiments. We also thank Dr. David Rowe, Li Chen and the rest of the components of Bonebase for the stain carried out on mouse samples.

