## supplementary appendix for "RANKL inhibition with denosumab improves fibrous dysplasia by decreasing lesional cell proliferation and increasing osteogenesis"

### Denosumab for Fibrous Dysplasia of Bone

List of investigators:

### Supplementary Methods

#### FD Mouse Model

At 10 weeks of age, expression of Gα_s_^R201C^ was induced in the limbs by switching to doxycycline-supplemented food (100 ppm Purina Mod LabDiet 5001, PMI Nutrition International, Saint Louis, MO). Six mice received anti-RANKL antibody (6 mg/Kg, BE0191, Bioxcell Lebanon, NH) and 6 received rat IgG2A isotype control, as well as 6 littermate controls not carrying tet- GsαR201C (6 mg/Kg, BE0089, Bioxcell) by subcutaneous injections on days 28, 30, 35, 42, 49 and 56 after induction. On day 58 all mice were euthanized, and FD or control tissue was extracted from the distal ulna and radius, cleaned from muscle and tendons, and snap-frozen for RNA analysis. Mice were then perfused with PBS and Z-fix fixative, and both hindlimbs were extracted for histology and µCT analysis.

#### Mouse plasma measurements

120 µL of mouse blood was collected weekly via retro-orbital eye collection. At the time of euthanasia, 500-1000 µL was obtained from the vena cava. Blood was stored in heparinized vials and plasma was obtained. TRAP5b, CT-X and P1NP was measured using IDS ELISA kits SB-TR103, AC-06F1 and AC-33F1, respectively.

#### Mouse X-rays and microCT

Mice were anesthetized with 2-5% of isoflurane and X-ray images of the hind limbs were obtained on a Faxitron Ultrafocus system (Hologic, Marlborough MA). A semi-quantitative score was developed and validated to quantify the disease burden (Suppl table 1). Three independent examiners blindly evaluated X-rays of both hindlimbs for each mouse and timepoint in an independent fashion.

Right hindlimbs were dissected and scanned using a Scanco µCT 50 at 10 µm, 70 kVp, 80 µA, and 900 ms integration time (Scanco, Wangen-Brüttisellen, Switzerland). Reconstructed images were analyzed with Analyze 14 (AnalyzeDirect, Overland Park KS) and calibrated against hydroxyapatite phantoms with known densities. The volume of interest (VOI) was defined as the distal tibia sector between 500 µm below the fibula insertion point and 200 µm above the intermedium. The Smart Trace tool was used to outline the distal tibia every 50 µm in the sagittal direction and then Propagate Objects was used to connect the 2D tracings, semi-automatically segmenting the VOI.

Following segmentation, the average bone mineral density (BMD) in milligrams of hydroxyapatite per cubic centimeter (mg HA/cm3) was obtained for the entire VOI. Distal tibiae were further analyzed by obtaining the number of voxels per BMD unit and binned into three categories: soft tissue (<350 mg HA/cm3), partially mineralized tissue (350-600 mg HA/cm3), and mineralized tissue (>600 mg HA/cm3).

#### Bone marrow explant and cell culture

Tibiae and femurs were dissected from uninduced mice or wild-type littermates and bone marrow cells (BMSCs) were pooled, plated and cultured as previously described without immunoselection ^1^. Cultures below passage 4 were used for the experiments. For GαsR201C expression induction in BMSCs subset of the explants, cells were plated at ~40% confluency in 6-well plates as well as in 24-well plates, and angiogenesis µ-slides (cat no. 81506 Ibidi, Firchburg, WI) and treated with 5µM doxycycline (Sigma, # D9891-5G). During induction, media were refreshed daily. Neutralizing RANKL antibody (BE0191, Bioxcell) was added at 1µg/mL.

MC3T3-E1 clone 4 and 14 cells were cultured as previously described^2^.

#### Histology and cell culture staining

##### Tissue and culture preparation

Dissected mouse hindlimbs and human baseline and post-treatment bone biopsies were fixed in Z-fix (Anatech, USA) overnight at 4°C. Samples for paraffin embedding (PE) were decalcified in 0.25 M EDTA at 4°C. Samples were then embedded and sectioned into 5 µm sections. Slides were stored in 4°C or -20°C until staining. Sections from all PE samples were deparaffinized in xylene and rehydrated using a graded ethanol series for subsequent staining. Tissue morphology was assessed by staining sections for H&E.

Tissue samples for cryosectioning were shipped to Bonebase (at UConn Health, Farmington, CT) in formalin and in wet ice, embedded and cryosectioned following the methods in Dyment et al ^3^.

Cell cultures were fixed with warm freshly prepared 4% formaldehyde in PBS (Sigma, F1268) at 37^◦^C.

##### TRAP and ALP enzymatic detection

Mouse cryosections were stained for TRAP and ALP enzymatic activity at Bonebase following previously described methods ^3^. Mouse culture TRAP staining was performed with Cosmo Bio LTD TRAP Staining Kit (Cat no. PMC-AK04F-COS, Cosmo Carlsbad, CA). Human PE sections were incubated with TRAP staining solution (Wako, Cat No 294-67001) for 25 min at RT, then counterstained with Methyl Green and mounted using EcoMount.

##### Immunohistochemistry

PE sections were deparaffinized in xylene and rehydrated using a graded ethanol series. Endogenous peroxidase activity was blocked using 3% H_2_O_2_ in methanol. For Runx2 and Mcm2, antigen retrieval was performed using Uni-Trieve (Innovex Biosciences, Cat No NB325) for 30 min at 55°C or 45 min at 70°C, respectively. Non-specific binding was blocked using goat or rabbit serum (Vector, Cat No PK-6105 and PK-6101) as appropriate. Next, both human and mouse sections were incubated with rabbit anti-Runx2 (1:400, Abcam, Cat No ab192259), rabbit anti-Mcm2 (1:200, Abcam, Cat No 108935), human sections were incubated with rabbit anti-osteocalcin (1:200, Proteintech, Cat No 23418-1-AP), and mouse sections were incubated with goat anti-sclerostin (1:100, R&D Systems, AF1589) overnight at 4°C. Rabbit and goat isotype control antibodies were used at a similar concentration (BioLegend, Cat No 910801; R&D Systems, Cat No AB-108-C, respectively) and no significant unspecific staining was observed (not shown). Sections were then incubated with corresponding goat anti-rabbit or rabbit anti-goat biotinylated secondary antibodies (Vector, Cat No PK-6105 and PK-6101) for 45 min at RT, followed by VECTASTAIN Elite ABC Reagent (Vector, Cat No PK-6100) for 30 min at RT. Finally, staining was developed using DAB-EASY tablets (Acros Organics, Thermo Fisher Scientific, Cat No AC328005000) until desired stain intensity was achieved. Samples were counterstained with Methyl Green (Vector, H-3402-500) and mounted using EcoMount (Biocare Medical, Cat No EM897L).

##### In vitro fluorescence and immunofluorescence

Mouse marrow explant cultures were stained with antibodies against Tsg101 (NOVUS, 4A10), Runx2 (Abcam, 76956), Ki-67 (CST, D3B5) and Rank (Abcam, 13918). Mouse-Alexa555 (CST, 4409) and Rabbit-Alexa647 (CST, 4414) were used as fluorescent secondary antibodies. Osteoclast fusion was evaluated by fluorescence microscopy as previously described ^4^ using phallodin-Alexa488 and Hoechst (Invitrogen, cat no. H3570 and A30106 respectively) to label actin cytoskeleton and nuclei. For all stains, cells were permeabilized 10 min in PBS with 0.1% Triton X100 in PBS and 5% FBS (IF Buffer) was used to suppress non-specific binding. Then the cells were incubated with primary antibodies overnight in IF Buffer. After washes in IF Buffer, we placed the cells for 2h at room temperature in IF Buffer with secondary antibodies. Images were captured on a Zeiss LSM 800 airyscan, confocal microscope using a C-Apochromat 63x/1.2 water immersion objective.

##### mRNA in situ hybridization

For human *SOST* mRNA hybridization, sections were deparaffinized and incubated with RNAscope Hydrogen Peroxide (Advanced Cell Diagnostics [ACD], Cat No 322335) for 10 min at RT. Target retrieval was performed using ACD Custom Pretreatment Reagent (ACD, Cat No 300040) for 45 min at 40°C. Probes were hybridized as instructed and consisted of dapB (bacterial gene, negative control), PPIB (housekeeping gene), and SOST (sclerostin). The remaining hybridization steps were performed using RNAscope 2.5 HD Detection Reagents- RED (ACD, Cat No 322360) with the exception of AMP5 incubation which was extended to 60 min. Sections were counterstained with 50% Gil’s Hematoxylin I (Sigma-Aldrich, Cat No GHS132) and mounted with EcoMount (Biocare Medical, Cat No EM897L).

In situ hybridization of Gα_s_ p.R201 variants mRNA was achieved using Advanced Cell Diagnostics [ACD] Basescope duplex system following manufacturer protocols. Briefly, human sections were incubated with RNAscope Hydrogen Peroxide (ACD, Cat No 322335) for 10 min at RT, retrieved with ACD Custom Pretreatment Reagent (ACD, Cat No 300040) for 60 min at 40°C. Positive controls PPIB (high-expression housekeeping gene) and POLR2A (low-expression housekeeping gene) and negative control dapB (bacterian gene) probes, and custom-designed probes for wild-type GNAS (ACD, Cat No 1061061-C1) as well as the two most common mutations in FD: GNASR201C (c.601C>T, ACD, Cat No 1061041-C2) and GNASR201H (c.602G>A, ACD, Cat No 1061051-C2). BaseScope Duplex Detection Reagent Kit (ACD, Cat No 323810); AMP7 and AMP11 times were increased to 45 min and 60 min, respectively, to increase staining intensity. Sections were counterstained using 50% Gil’s Hematoxylin I and mounted with VectaMount Permanent Mounting Medium (Vector, Cat No H-5000).

##### Microscopy imaging

Chromogenically stained PE sections were scanned using a NanoZoomer S60 Digital slide scanner (Hamamatsu, Cat No C13210-01) at 400X magnification.

Fluorescent cryogenic sections were scanned using a Axioscan 7 fluorescence scanner (Zeiss, Oberkochen, Germany) ^3^ with filters appropriate for the detection of DAPI (Em 460nm, Ab 350nm), Elf97 TRAP (Em 550nm, Ab 375nm) and ALP (Em 605nm, Ab 545nm). Scanned slides were manually aligned and converted to Adobe Photoshop .psd files.

Cell immunofluorescence images were captured on a Zeiss LSM 800 airyscan, confocal microscope using a C-Apochromat 63x/1.2 water immersion objective. For osteoclast fusion assay, 8 randomly selected fields of view were imaged using Alexa488, Hoechst and phase contrast compatible filter sets (BioTek) on a Lionheart FX microscope using a 10x/0.3 NA Plan Fluorite WD objective lense (BioTek) using Gen3.10 software (BioTek).

##### Microscopy quantification

For bone content and cellularity, images from H&E-stained PE sections were analyzed using a custom script in ImageJ (NIH). For human sections, complete biopsy sections were analyzed. For mice, 1-3 fields of 0.6 by 0.4 mm of randomly selected FD lesional tissue were analyzed per sample. Non-bone or non-FD tissue were removed using Adobe Photoshop 2021 when necessary (Adobe, San Jose, CA). On image J, pixel to area calibration was performed and mineralized and fibrous tissue areas were manually traced, and nuclei were selected in these areas using a semi-automatic color threshold selection followed by a watershed separation plugin. Nuclei number and mineralized and fibrotic areas were calculated using the “analyze particles” function. Total nuclei number obtained with this method was used as denominator to calculate the ratio of positive cells for TRAP activity, Rank, and Mcm2 in consecutive sections form the human biopsies.

For mouse TRAP and ALP analysis in mouse FD cryosections, multiplexed fluorescence 1 by 1 mm images of the distal tibiae were opened in Adobe Photoshop and color levels were adjusted and exported as single-layer tiff files corresponding to TRAP, ALP or DAPI for image quantification. Images were then analyzed with QuPath version 0.3.2, an open-source software for digital pathology analysis ^5^. After calibrating the pixel to area ratio of the images, cell nuclei were identified by DAPI staining and counted using the “cell detection” function. TRAP and ALP-stained areas were traced using the “pixel classification” function in which the area of fluorescence (µm2) is calculated based on a pixel color intensity threshold. Average TRAP and ALP stain per cell was calculated as the ratio of µm2 TRAP or ALP positive area and number of nuclei in each quantified image.

We used QuPath point tool in PE scans to manually label and count cells positive for TRAP, Rank (multinuclear osteoclasts or mononuclear precursors, Mcm2 and, sclerostin-stained (either by RNA hybridization in humans or immunohistochemistry in mice) and non-stained osteocytes. For ACD Basescope™ quantification, we used this tool in 400x images to label hematoxylin-stained nuclei in proximity of detected Gα_s_^R201C^ or Gα_s_^R201H^ mRNA molecules (green dots), quantified as mutant cells, regardless of the neighboring presence of wildtype Gα_s_ mRNA molecules (red dots). Nuclei only associated to wildtype Gα_s_ mRNA molecules where quantified as wildtype cells. Positive controls PPIB (high-expression housekeeping gene) and POLR2A (low-expression housekeeping gene) and negative control dapB (bacterian gene) probes were used.

Osteoclast fusion efficiency was evaluated as the number of fusion events between osteoclasts with obvious ruffled boarders and ≥3 nuclei in phalloidin-Alexa488/Hoechst images, as described previously ^6^. Since regardless of the sequence of fusion events, the number of cell-to-cell fusion events required to generate syncytium with N nuclei is always equal to N-1, we calculated the fusion number index as Σ (Ni − 1) = Ntotal − Nsyn, where Ni = the number of nuclei in individual syncytia and Nsyn = the total number of syncytia.

All assessments that required human evaluation were examined by at least 3 trained researchers in a blinded fashion.

##### In vitro bone resorption assay

Bone resorption was evaluated using bone resorption assay kit from Cosmo Bio USA according to the manufacturer’s instructions. In short, fluoresceinamine-labeled chondroitin sulfate was used to label 24-well, calcium phosphate-coated plates. Media were collected at 4-5 days post induction and fluorescence of the media was evaluated as recommended by the manufacturer.

##### Enrichment and quantification of Extracellular Vesicle (EV) Fractions

*Ex vivo* cultures were cultured in complete culture media supplemented with FBS depleted of EVs (via ultracentrifugation at 150,000xg for >2 hours). 24 hours later, conditioned media was collected, and cells/large cell debris were depleted via centrifugation (15 mins at 4,000xg). Next, an EV fraction was enriched via ultracentrifugation (150,000xg for 1.5 hours). Alternatively, exoEasy Maxi Kit (Qiagen, Hilden Germany) was used to isolate EVs. EV-enriched fractions were evaluated via Western Blot using anti-Tsg101 (cat no. 4A10, NOVUS, Louis MO) and anti-RANK (cat no. 13918 Abcam, Cambridge UK) antibodies. Tsg101 and Rank bands staining was quantified using densitometry via ImageJ.

##### RNA extraction and sequencing

Mouse and human samples were snap-frozen, pulverized using an automated dry pulverizer (CP02 cryoPREP, Covaris, Woburn MA) and immediately transferred to Trizol (Thermofisher). Phenol-chloroform RNA extraction was carried out. cDNA synthesis, library construction and sequencing were performing by Novogene (University of California, Davis, CA). Briefly, messenger RNA was purified from total RNA using poly-T oligo-attached magnetic beads. After fragmentation, the first strand cDNA was synthesized using random hexamer primers followed by the second strand cDNA synthesis, end repair, A-tailing, adapter ligation, size selection, amplification, and purification. The library was checked with Qubit and real-time PCR for quantification and bioanalyzer for size distribution detection. Quantified libraries were pooled and sequenced on an Illumina NovaSeq 6000 sequencer, according to effective library concentration and data amount using 150 x 150 paired-end mode. After sequencing, the base-called demultiplexed (fastq) read qualities were determined using FastQC (v0.11.2) ^7^, aligned to the GENCODE v32 human genome (GRCh38.v32) form human samples and GENCODE M23 mouse genome (GRCm38.M23) for mouse samples and gene counts generated using STAR (v2.7.3a) ^8^. Post-alignment qualities were generated with Picard tools. An expression matrix of raw gene counts was generated using R ^9^ and filtered to remove low counts genes (defined as those with less than 5 reads in at least one sample). The filtered expression matrix was used to generate a list of differentially expressed genes between the sample groups using DESeq2 ^10^ and analyzed with principal component analysis (PCA). Reads corresponding to locations chr20:909,365 and chr20:58,909,366 were analyzed in order to detect and quantify GNAS p.R201C (C>T) and pR201H (G>A) substitutions respectively. An FD tissue genetic signature comprising 276 genes was derived from comparing WT versus Gnas R201C expressing mice that were more than ±2 log2 fold change and were significantly (adjusted p-value ≤ 0.05) differentially expressed between both groups. This gene signature was converted to human genes based on homology scores using the Biomart service, which reduced it to 202 genes. Likewise, a list of genes positively correlated with the proliferation marker PCNA in healthy human tissues was compiled from Venet et al ^11^. A custom .gmt file was compiled from the FD, meta PCNA signatures and MSigDB GO-derived genesets corresponding to osteoclast and osteoblast differentiation and activity biological processes. For osteoblasts, the geneset GO:0001649 “osteoblast differentiation” list was used. For osteoclasts, we combined two genesets: GO:0030316 “osteoclast differentiation” and GO:0045453 “bone resorption”. The gene set variation analysis (GSVA) ^12^ method was used to compute enrichment scores for each sample against these selected genesets using the Poisson kernel and the resulting matrix was submitted to unsupervised clustering using the euclidean metric. Selected gene expression heatmaps were generated from log transformed TMM-normalized gene counts using the heatmap.2 R function.

### Supplementary tables

#### Table S1: Eligibility criteria

| Inclusion Criteria | Exclusion Criteria |
| --- | --- |
| - Age > or equal to 18 years - Skeletal maturity with fusion of epiphyses documented radiographically - Concomitant enrollment in the companion Screening and Natural History protocol 98-D-0145 - Confirmed diagnosis of Fibrous Dysplasia - Self-reported FD-related bone pain at a site of FD, present for at least 3 months - Ability to understand and provide informed consent - Willing and able to complete the protocol scheduled assessments and medication regimen - Willing and able to take calcium and vitamin D supplements provided by NIH - Women of child bearing potential must use two forms of acceptable contraception: hormonal contraception (birth control pills, injected hormones or vaginal ring); intrauterine device and/or barrier methods (condom or diaphragm) used with spermicide. Surgical sterilization (hysterectomy, tubal ligation, or vasectomy in a partner) is considered one form of contraception therefore only one other form of contraception will be required in these subjects. Verbal confirmation will be obtained at screening that two forms of acceptable contraception are being used, and that the subjects agree to use the two forms of contraception from the time they sign study consent through five months after study completion (19 months total). - Serum calcium or albumin-adjusted serum calcium within the normal range for the NIH laboratory. | - Administration of denosumab within the previous year - Use of bisphosphonates within one year prior to first day of the denosumab administration (Day 0). Examples of bisphosphonates include: pamidronate (Aredia), Alendronate (Fosamax) and Zoledronate (Zometa). - Prior history, or current evidence, of osteomyelitis/osteonecrosis of the jaw or presence of an active dental or jaw condition which requires oral surgery - Planned invasive dental procedure for the course of the study - Presence of non-healed dental or oral surgery - Orthopedic procedure performed less than 12-weeks prior to first day of the denosumab administration (Day 0) - Acute fracture less than 12-weeks prior to first day of the denosumab administration (Day 0) - 25-hydroxyvitamin D level than 30 ng/mL (patients will be eligible for re-screening after a repletion period lasting no longer than 6 months) - Untreated or inadequately treated hypophosphatemia, defined as serum phosphate levels below the NIH normal range (patients will be eligible for re-screening after initiation or optimization of phosphorus replacement no longer than 6 months) - Subject is pregnant or breast feeding, or planning to become pregnant or breastfeed while on study and through 5 months after completion of treatment - Use of another investigational agent within the last 3 months prior to the first day of the denosumab administration (Day 0) - Subject has known sensitivity to any of the products to be administered during the study (e.g. polyketide antibiotics, mammalian derived products, calcium, or vitamin D) |

#### Table S2: Pulmonary functions tests in response to denosumab for subject DB02

|  | Baseline  (pre-denosumab) | 6-months  (post-denosumab) | 14-months  (8-months after denosumab discontinuation) | 26-months  (12-months after denosumab restart)^1^ |
| --- | --- | --- | --- | --- |
| FEV1 (% ref) | 38 | 48 | 32 | 36 |
| FVC (% ref) | 34 | 43 | 30 | 35 |
| FEV1/FVC | 87 | 86 | 83 | 82 |
| TLC (% ref) | 45 | 51 | 39 | 49 |
| DLCO (% ref) | 29 | 34 | 34 | 37 |
| Supplemental oxygen requirement | 2 liters/minute | none | 2 liters/minute | none |
| ^1^Based on declining pulmonary function at the 14-month timepoint, clinically indicated denosumab was restarted at a dose of 120 mg every 12 weeks | | | | |

#### Table S3: Bone turnover markers during and after denosumab treatment

| Timepoint | DB01 | DB02 | DB03 | DB04 | DB05 | DB07 | DB08 | DB09 |
| --- | --- | --- | --- | --- | --- | --- | --- | --- |
| C-Telopeptides (normal range 25-573 pg/mL) | | | | | | | | |
| Baseline | 525 | 2011 | 742 | 800 | 800 | 568 | 1346 | 1357 |
| 6-months | 80 | 772 | 157 | 114 | 106 | 67 | 134 | 142 |
| 9-months | 69 | 1692 | >5000^¶^ | 68 | 88 | 83 | 140 | 235 |
| 11-months | n/a | 1973 | 630 | 127 | 274 | 66 | 887 | 1494*, 2558^ⴕ¶^ |
| 14-months | 162 | 1051 | 1022 | 378 | 1212 | 853^¶^ | 1478 | 1513 |
| Procollagen 1 Propeptide (normal range 16-83 mcg/L) | | | | | | | | |
| Baseline | 123 | 570 | 1197 | 288 | 267 | 195 | 237 | 780 |
| 6-months | 16 | 53 | 95 | 21 | 26 | 26 | 33 | 40 |
| 9-months | 19 | 230 | 1308^¶^ | 18 | 17 | 29 | 34 | 45 |
| 11-months | n/a | 236 | 191 | 25 | 57 | 15 | 106 | 160*, 866 ^§¶^ |
| 14-months | 16 | 160 | 530 | 116 | 294 | 179^¶^ | 227 | 508 |
| Interventions | ZA (6-mo) | ZA (6-mo) | ∘ ZA (6-mo)  ∘ ZA, IV fluids, calcitonin, dmab 60 mg (9-mo) | ZA (6-mo) | ZA (6-mo, 11-mo, 14-mo) | ZA (6-mo, 9-mo) | ZA (6-mo, 9-mo) | ZA (6-mo, 9-mo, 11.5-mo) |

Timepoints in this table were chosen to encompass start of therapy (baseline), completion of therapy (6-months), peak bone turnover values, and completion of the post-discontinuation monitoring (14-months, i.e. 8-months after denosumab discontinuation). n/a = data not available, ZA = zoledronic acid, IV = intravenous fluids, dmab = denosumab, mo = month. ^¶^indicates hypercalcemia at this timepoint. *indicates 11-month value. ^ⴕ^indicates peak value, which occurred at 11.5-months. ^§^ indicates peak value, which occurred at 12.5-months.

#### Table S4: FD tissue genetic signature

|  | **Human**  **Biopsy** | **Mouse distal ulna** | |  |  | **Human**  **Biopsy** | **Mouse distal ulna** | |
| --- | --- | --- | --- | --- | --- | --- | --- | --- |
| **Gene** | **6 month vs baseline** | **FD+****αRANKL vs FD** | **FD vs WT** |  | **Gene** | **6 month vs baseline** | **FD+αRANKL vs FD** | **FD vs WT** |
| SIGLEC15 | -12.1 | -4.1 | 2.4 |  | COL5A2 | -1.7 | -2.2 | 2.1 |
| DCSTAMP | -9.1 | -4.8 | 2.7 |  | ITGAV | -1.7 | -2.1 | 2.1 |
| ATP6V0D2 | -8.4 | -3.2 | 2.0 |  | COL1A1 | -1.6 | -3.4 | 2.4 |
| GAL | -6.5 | -4.5 | 4.0 |  | COL6A3 | -1.6 | -1.4 | 2.5 |
| CPZ | -4.4 | -3.8 | 2.4 |  | CGREF1 | -1.6 | -3.3 | 2.4 |
| SLC9B1 | -4.0 | -3.1 | 2.8 |  | COL5A1 | -1.5 | -2.3 | 2.4 |
| SLC37A2 | -3.8 | -3.5 | 2.4 |  | COL1A2 | -1.4 | -3.0 | 2.2 |
| SLC9B2 | -3.4 | -4.7 | 2.7 |  | PRSS35 | -1.4 | -3.6 | 3.4 |
| CDCP1 | -3.0 | ns | -2.4 |  | CREB3L1 | -1.4 | -2.4 | 2.2 |
| HAS2 | -2.6 | -2.1 | 2.4 |  | GXYLT2 | -1.3 | -2.1 | 2.1 |
| SHTN1 | -2.6 | -2.3 | 2.1 |  | ALDH1L2 | -1.3 | -3.5 | 2.6 |
| CTHRC1 | -2.4 | -3.4 | 2.9 |  | MMP14 | -1.1 | -1.9 | 2.0 |
| FLG | -2.3 | 21.1 | -22.7 |  | C1QTNF6 | -1.1 | -2.4 | 2.3 |
| CKB | -2.1 | -3.5 | 2.9 |  | ZNF469 | -1.1 | -2.2 | 2.2 |
| CTSK | -2.1 | -3.9 | 2.7 |  | COL12A1 | -1.0 | -1.3 | 2.0 |
| GALNT5 | -1.9 | -2.0 | 2.1 |  | TUBB6 | -0.7 | -2.3 | 2.2 |
| MFAP2 | -1.8 | -1.3 | 2.1 |  | CA4 | 4.9 | ns | 2.2 |
| COL11A1 | -1.7 | -2.8 | 2.1 |  | MYOZ2 | 9.5 | 1.4 | -4.5 |

FD signature genes significantly regulated by denosumab in human FD tissue (left column). Ortholog mouse genes in αRANKL-treated mouse FD tissue showed overall similar regulation in (central column). Mouse FD tissue presented an opposite regulation in comparison to WT bone (left column). Genes with at least 2-fold change and p<0.05 were selected in all groups, and fold-changes are shown in Log_2_ base.

| **Score 🡪**  **Bone(s) ↓** | **1** | **2** | **3** | **4** | **5** | **6** |
| --- | --- | --- | --- | --- | --- | --- |
| **Femur** | Focal lytic areas covering <10% of the femur Typically just one spot in the distal metaphysis. | Extended lytic areas covering 10-25% of the femur.  OR Focal sclerotic only covering <10% of the femur and immediately proximal to the distal metaphysis | Lytic or lytic-sclerotic changes, covering 25-50% of the femur No Expansion | Lytic or lytic-sclerotic changes, covering >50% of the femur No Expansion | Lytic or lytic-sclerotic changes, covering >50% of the femur Mild Expansion, diaphysis width <125% control | Lytic or lytic-sclerotic changes, covering >50% of the femur Severe Expansion, diaphysis width >125% control |
| **Tibia-fibula** | Focal lytic areas covering <10% of the tibia Typically just one spot in the distal submetaphysis. Rarely more proximal in the dyaphisis or in the tibial crest | Extended lytic areas covering 10-25% of the tibia No Expansion Mid-diaphisis unaffected | Lytic/sclerotic areas covering 25-50% of the tibia No Expansion Mid-diaphisis unaffected Typically no lesioned tissue proximal to the fibula insertion | Lytic/sclerotic areas covering 50-75% of the tibia with mild bowing (score 4a) OR lytic/sclerotic areas covering 25-50% of the tibia with focal expansion <150% in size (score 4b) | Lytic/sclerotic changes, covering 50-75% % of the tibia Focal Expansion - width >150% in size | Lytic/sclerotic changes, covering over 75% of the tibia Spread Expansion - width >150% normal |
| **Calcaneus** | Focal lytic areas No expansion | Extended lytic or lytic/sclerotic areas No expansion | Expansion Size 100-125% normal Lytic/sclerotic | Expansion Size >125% normal No ankylosis of the ankle Lytic/sclerotic | Expansion Size >125% normal Partial ankylosis of the ankle Lytic/sclerotic | Expansion Size >125% normal Complete ankylosis of the ankle Lytic/sclerotic |
| **Metatarsals** | Focal lytic areas in some metatarsals, immediately proximal to the distal metaphysis. It can be differentiated from a low-capture X-ray because in the radiolucent cortex is restricted to these areas (not continuous) | Extended lytic areas in some of the metatarsals. No Expansion | Expansion Width of affected bones < 150% Lytic and sclerotic | Expansion Width of affected bones 150-200% Unaffected areas in at least 1 metatarsal Lytic and sclerotic | Expansion Width of affected bones >200% Unaffected areas in at least 1 metatarsal Lytic and sclerotic | Expansion Width of affected bones >200% No unaffected areas Lytic and sclerotic |

#### Table S5: Mouse X-ray disease burden score

**Mouse disease burden score = (left femur + left tibia-fibula + left calcaneus + left metatarsals + right femur + right tibia-fibula + right calcaneus + right metatarsals) / 8**

### Supplementary figures

#### Figure S1 – Clinical flowchart

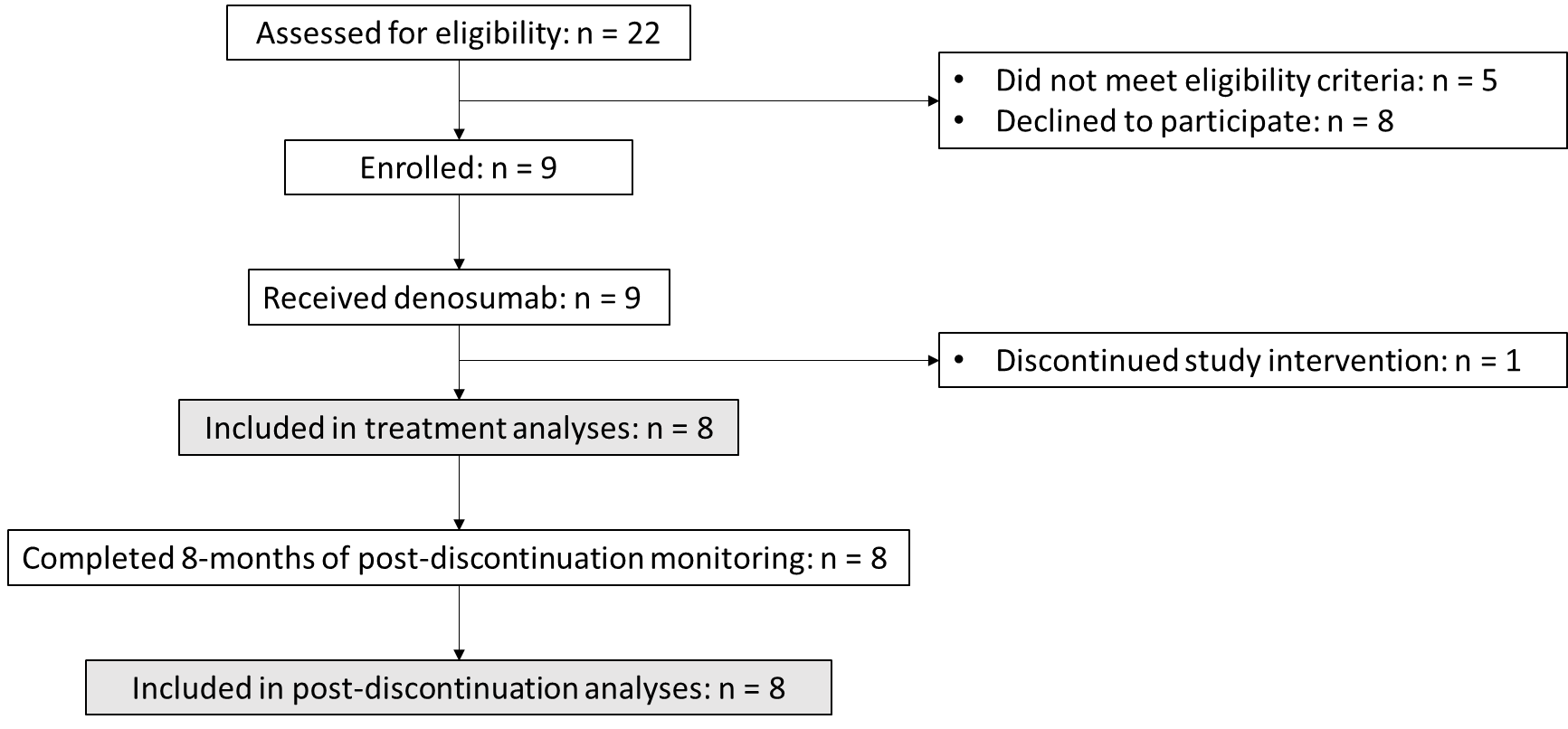

#### Figure S2 – RANK, OCN and RUNX2 immunostaining and quantification in human biopsies

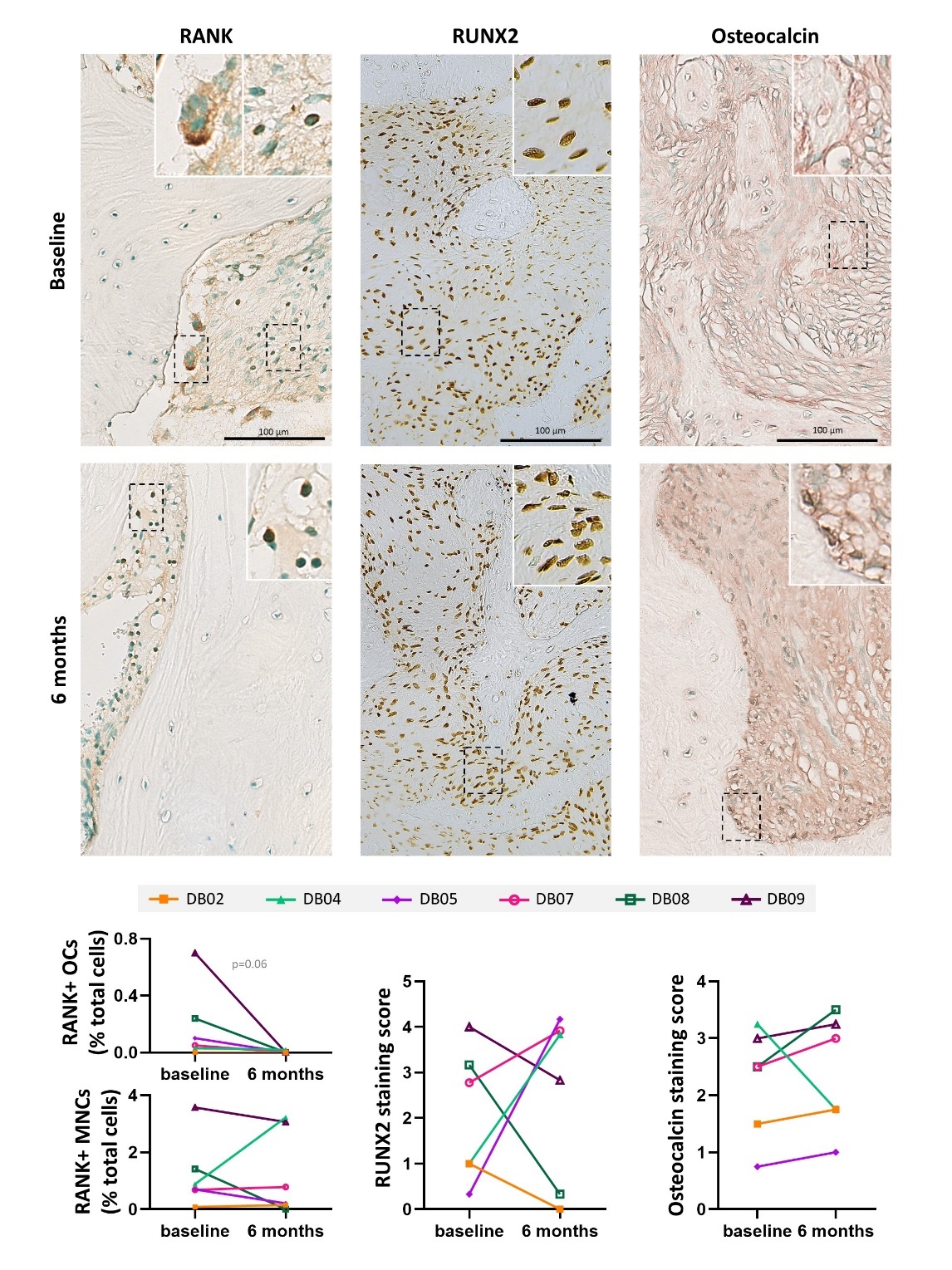

Figure S2 – Immunostaining and quantification of RANK, RUNX2 and Osteocalcin in human FD biopsies before and after 6 months of denosumab treatment. Clearance of RANK+ multinucleated cells was observed after denosumab treatment, although RANK+ mononuclear cells were still present after treatment. No change in the expression level of RUNX2, which had broad baseline expression within the fibrotic tissue. 5/6 biopsies presented mild non-significant increases in osteocalcin staining.

#### Figure S3 – H&E stains of biopsies before and after denosumab treatment

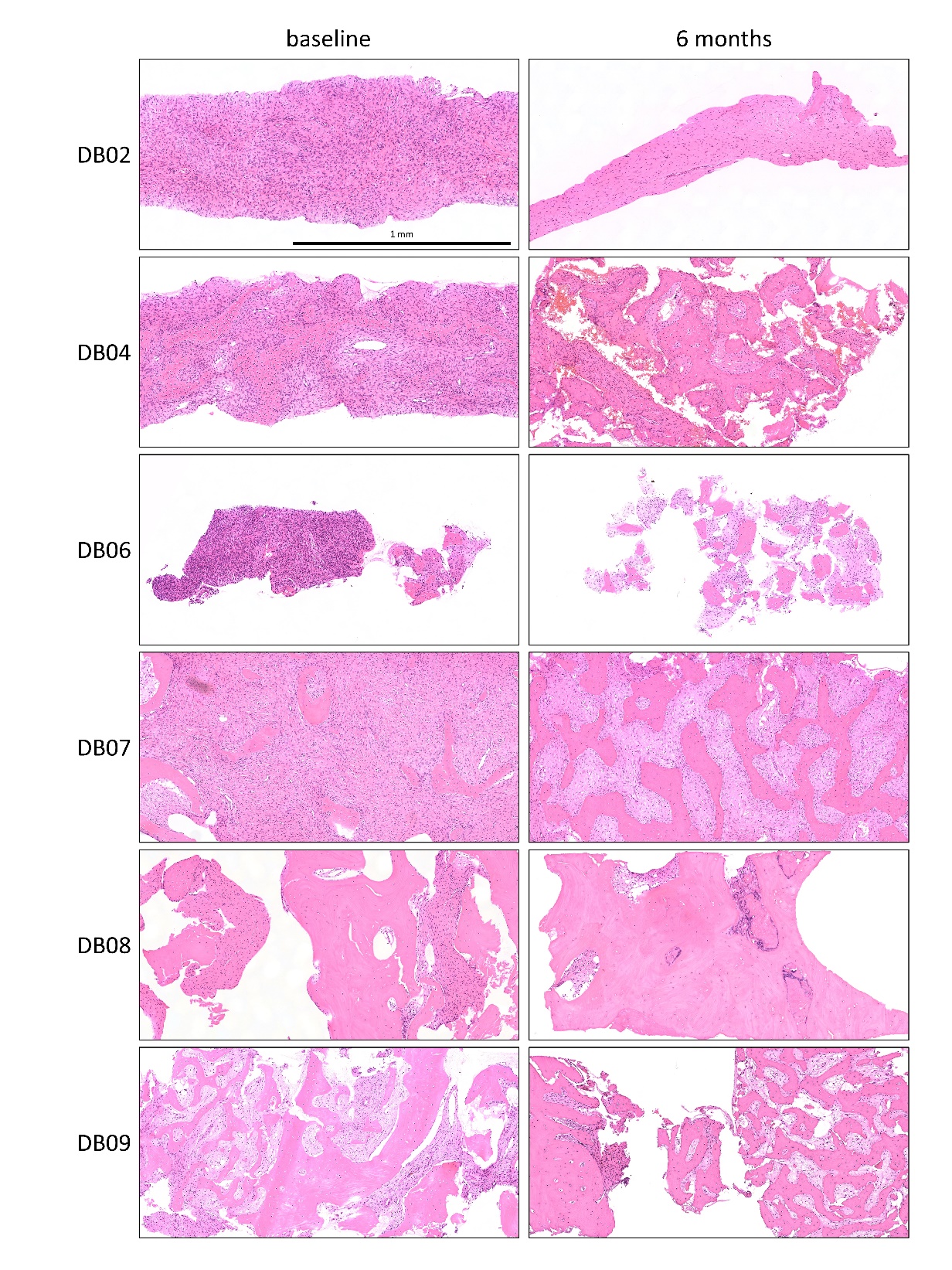

Figure S3. Low magnification images of a cross-section of all biopsies obtained from patients at baseline and 6 months post-treatment.

#### Figure S4 – ACD Basescope™ negative and positive controls

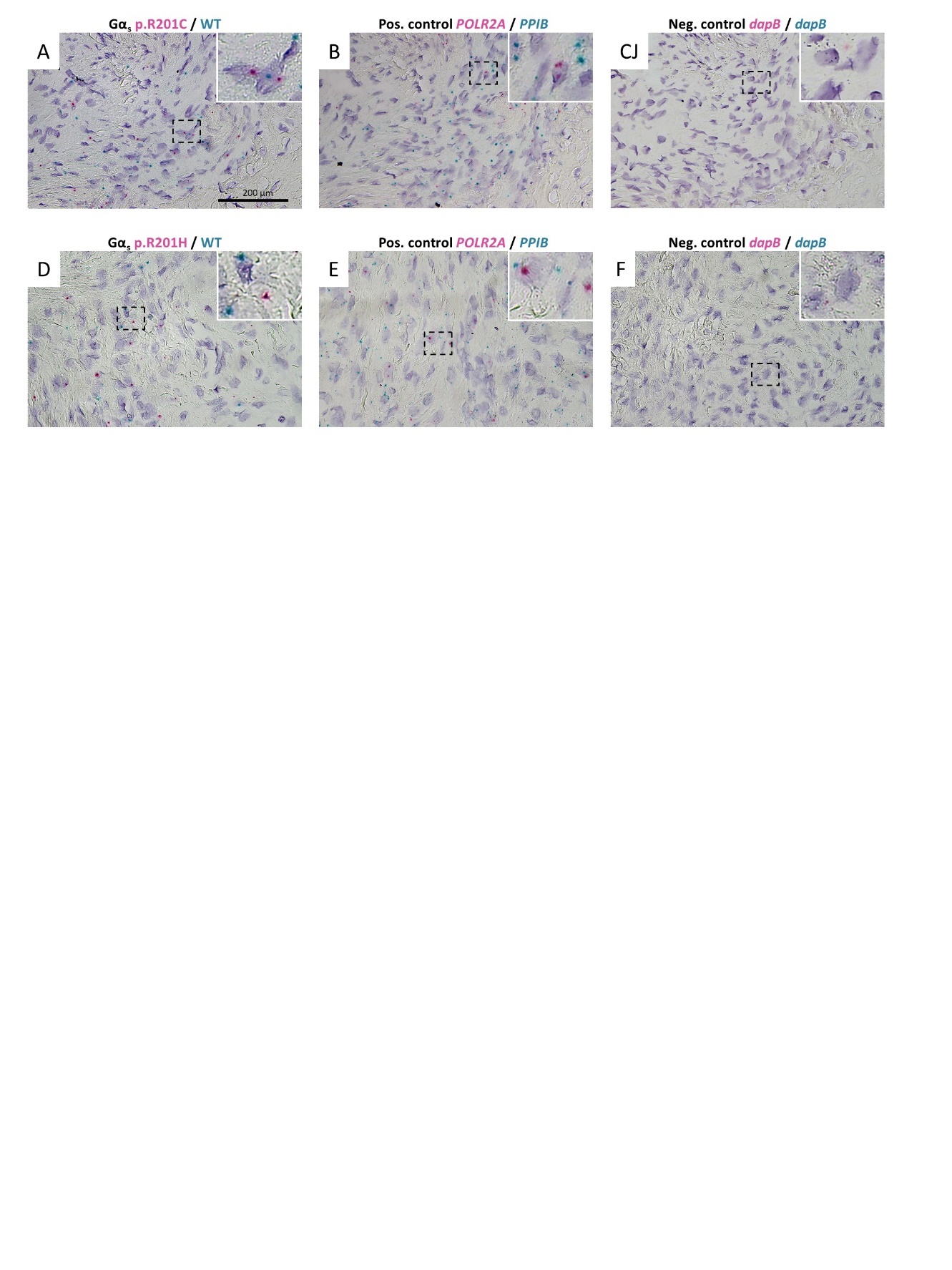

Figure S4. ACD Basescope™ stains showing the same region of a patient with Gα_s_ p.R201C (DB04, A-C) with Gα_s_ p.R201H (DB02, D-F) stained for Gα_s_ wildtype (green dots) and p.R201C/H variants (red dots) respectively (A, D); positive controls PPIB (high expression, geen dots in B and E) and POLR2A (low expression, red dots in B and E); and negative control dapB (red stain, undetectable in C and F).

#### Figure S5 – Selected gene expression heatmaps of human biopsies

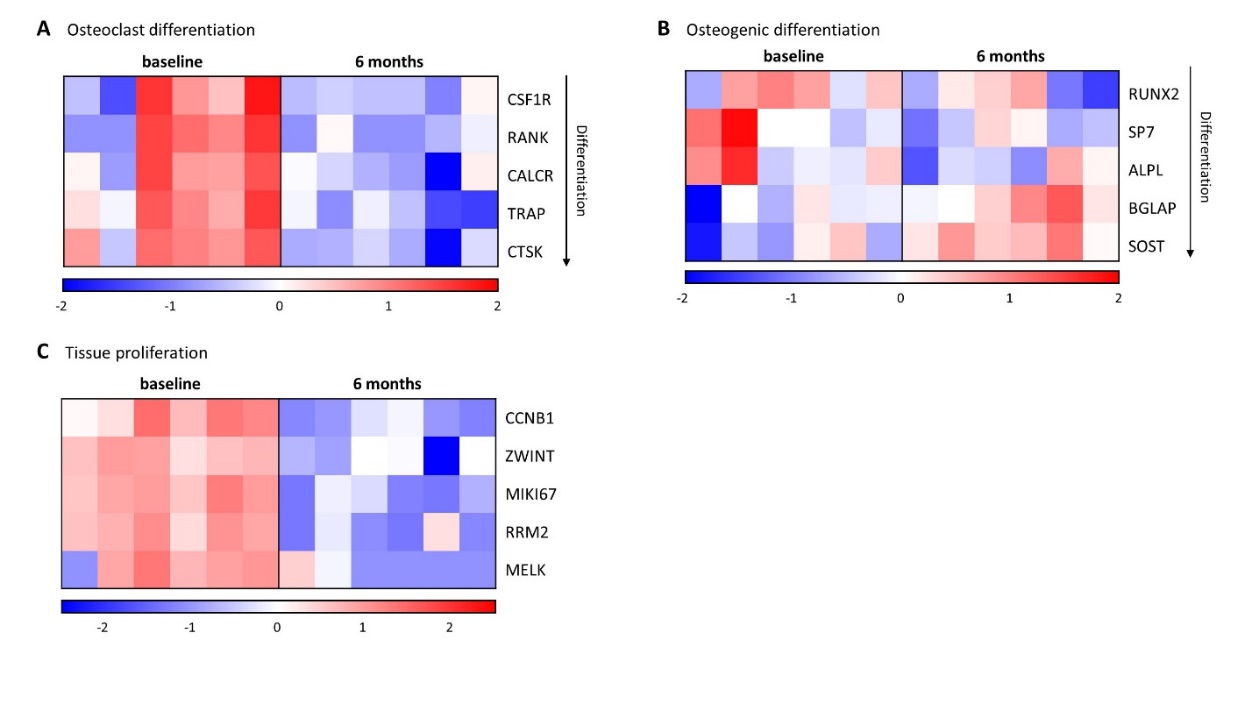

Fig S5. Heatmaps of selected, well characterized gene markers of different stages in osteclastic differentiation, osteoblastic differentiation and tissue proliferation. While all genes related to osteoclastic differentiation where consistently downregulated, early markers of osteoblastic differentiation and activity failed to show significant differences (RUNX2, SP7 and ALPL), late differentiation marker BGLAP (osteocalcin) and osteocyte terminal differentiation marker SOST were significantly upregulated after denosumab. Tissue proliferation markers were selected amongst the top 10 genes most co-regulated with PCNA in a broad collection of human tissue ^11^.

#### Figure S6 – µCT density categorization and average bone mineral density in FD tissue

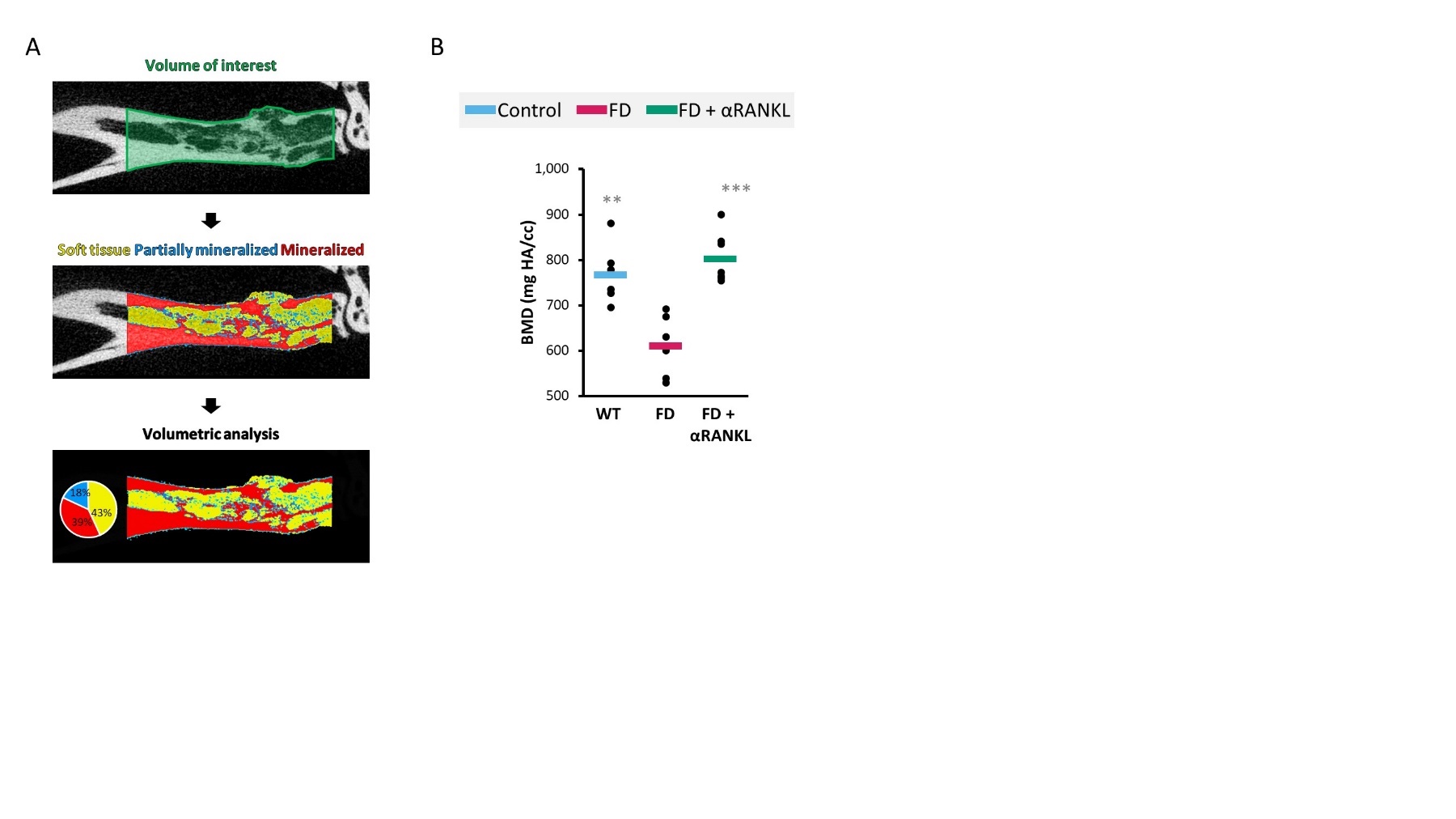

Figure S6. A) Method used for analyzing, partially mineralized and mineralized tissue in distal tibia FD lesions. Volume of interest was defined as the sector of the tibia between 500 µm below the fibula insertion point and 200 µm above the intermedium. Tissue density thresholds where applied as defined in fig 3B of the manuscript and the proportional abundancy of soft, partially mineralized, and mineralized volumes where calculated. B) Average bone mineral density of the complete volume of interest. Data is shown as individual mice (dots) and group averages (bars). ** is p<0.01 and *** is p<0.001 vs FD group.

#### Figure S7 – Mouse FD lesion progression and scoring examples

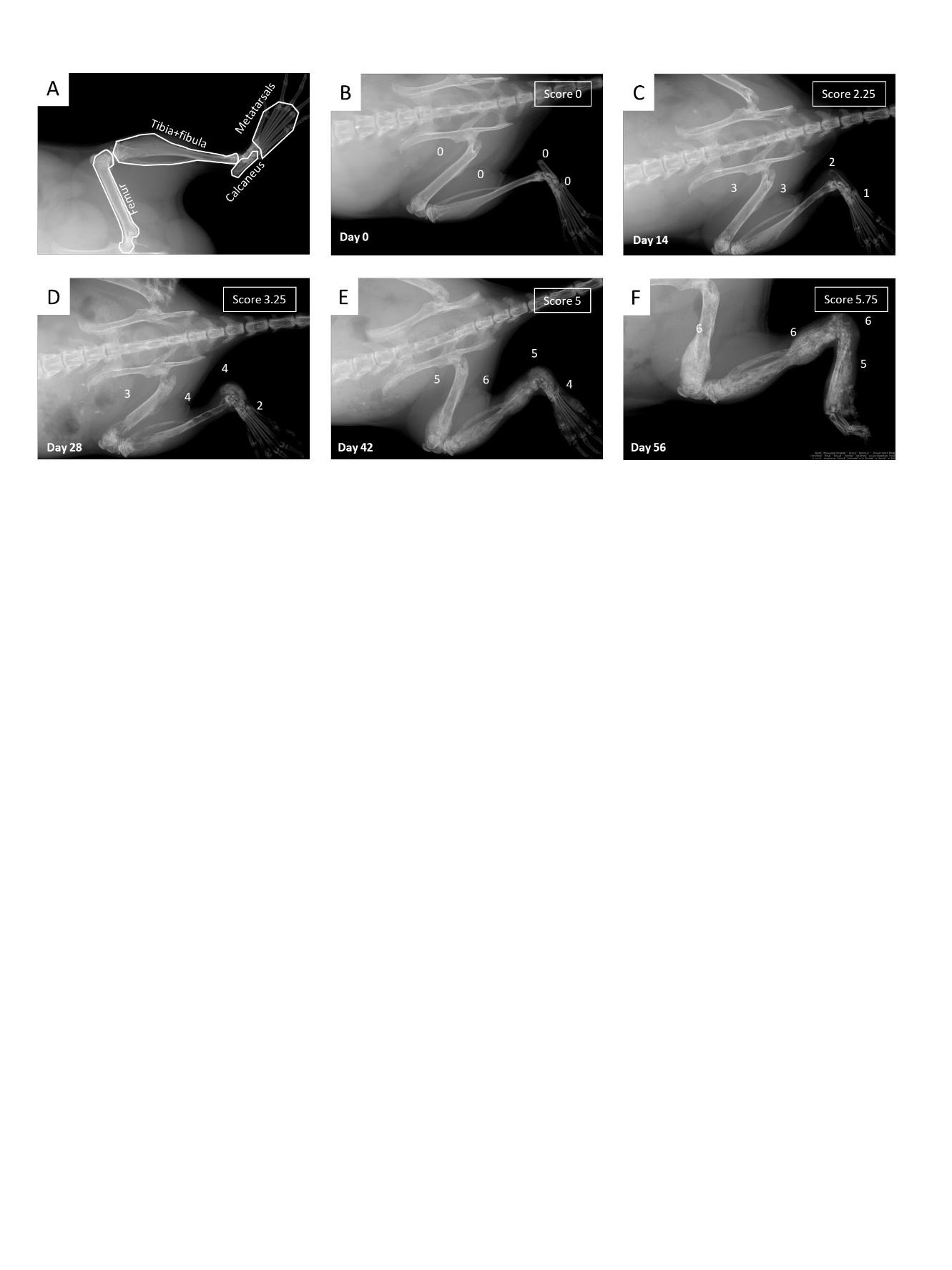

Figure S7 – Longitudinal progression of a FD mouse hindlimb lesion and its score. A) Hindlimb sectors receiving a score. B-F) Scores of each hindlimb sector along 56 days of disease progression.

#### Figure S8 – Mouse FD lesions H&E stains, bone content and cellularity

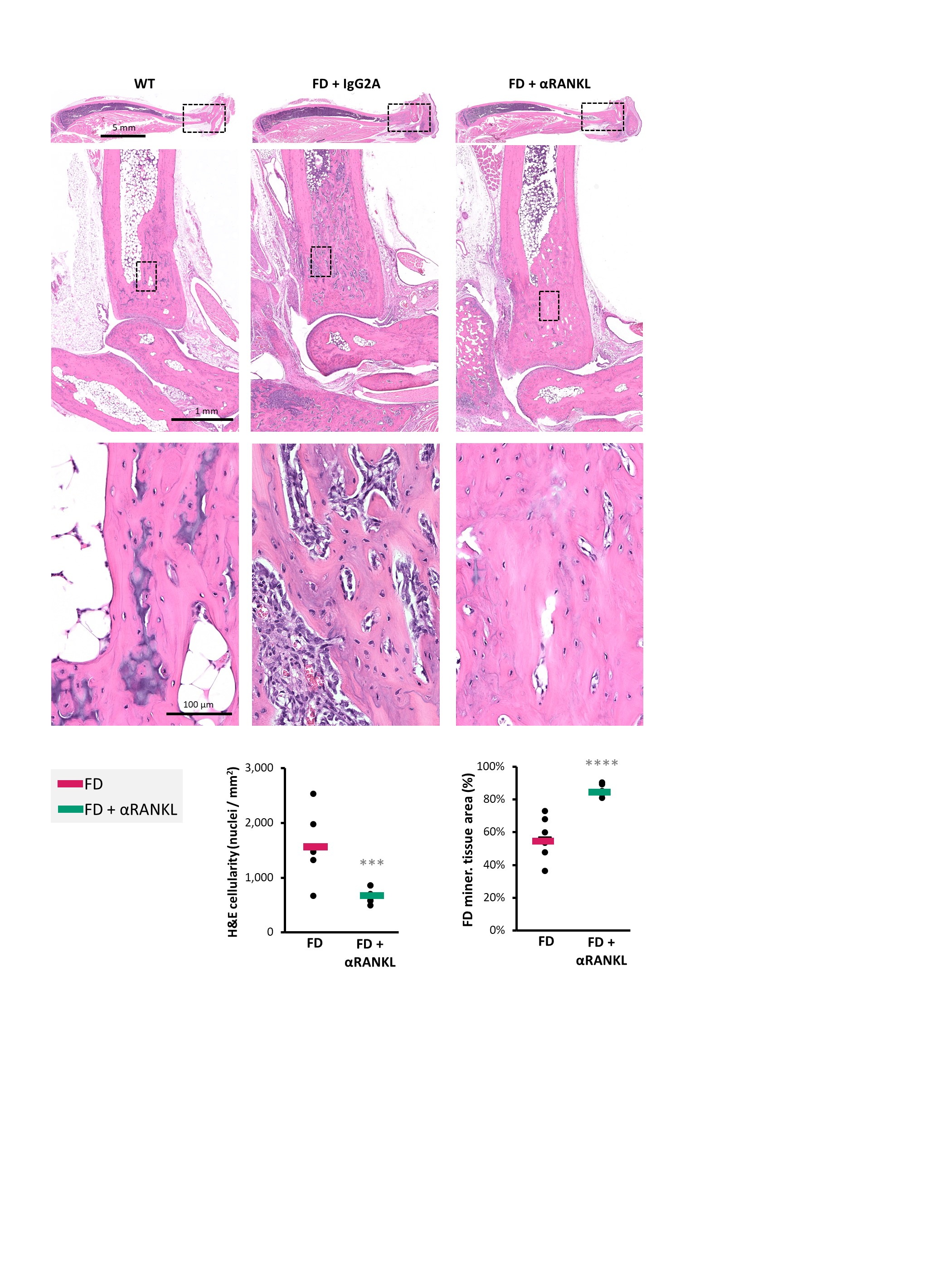

Figure S8 – H&E stains of WT and FD mice distal tibia and analysis of cellularity and bone content (mineralized tissue area) of the lesioned areas with or without αRANKL treatment. Data is shown as individual mice (dots) and group averages (bars). *** is p<0.001 and **** is p<0.0001 vs FD group.

#### Figure S9 – Mouse FD lesions enzyme histochemistry and immunohistochemistry

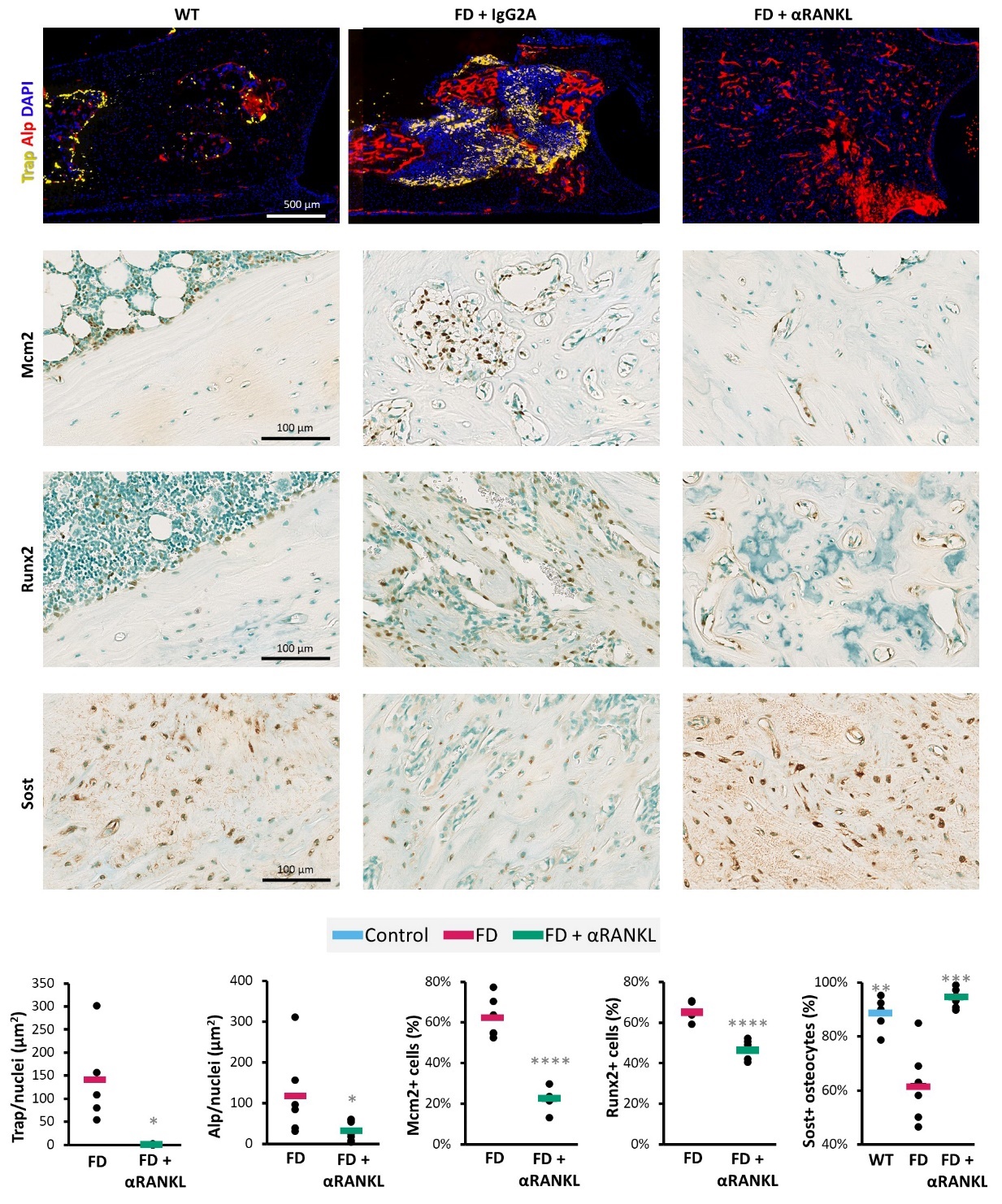

Figure S9: Trap, Alp enzymatic activity and DAPI nuclear staining; Mcm2, Runx2 and Sost immunostaining of WT and treated and untreated FD mice distal tibiae. Control distal tibia cortical bone was used to calculate the physiological level of Sost positivity within osteocytes. Data is shown as individual mice (dots) and group averages (bars). * is p<0.05, ** is p<0.01, *** is p<0.001 and **** is p<0.0001 vs FD group.

#### Figure S10 – Transcriptomic analysis of mouse FD lesional tissue

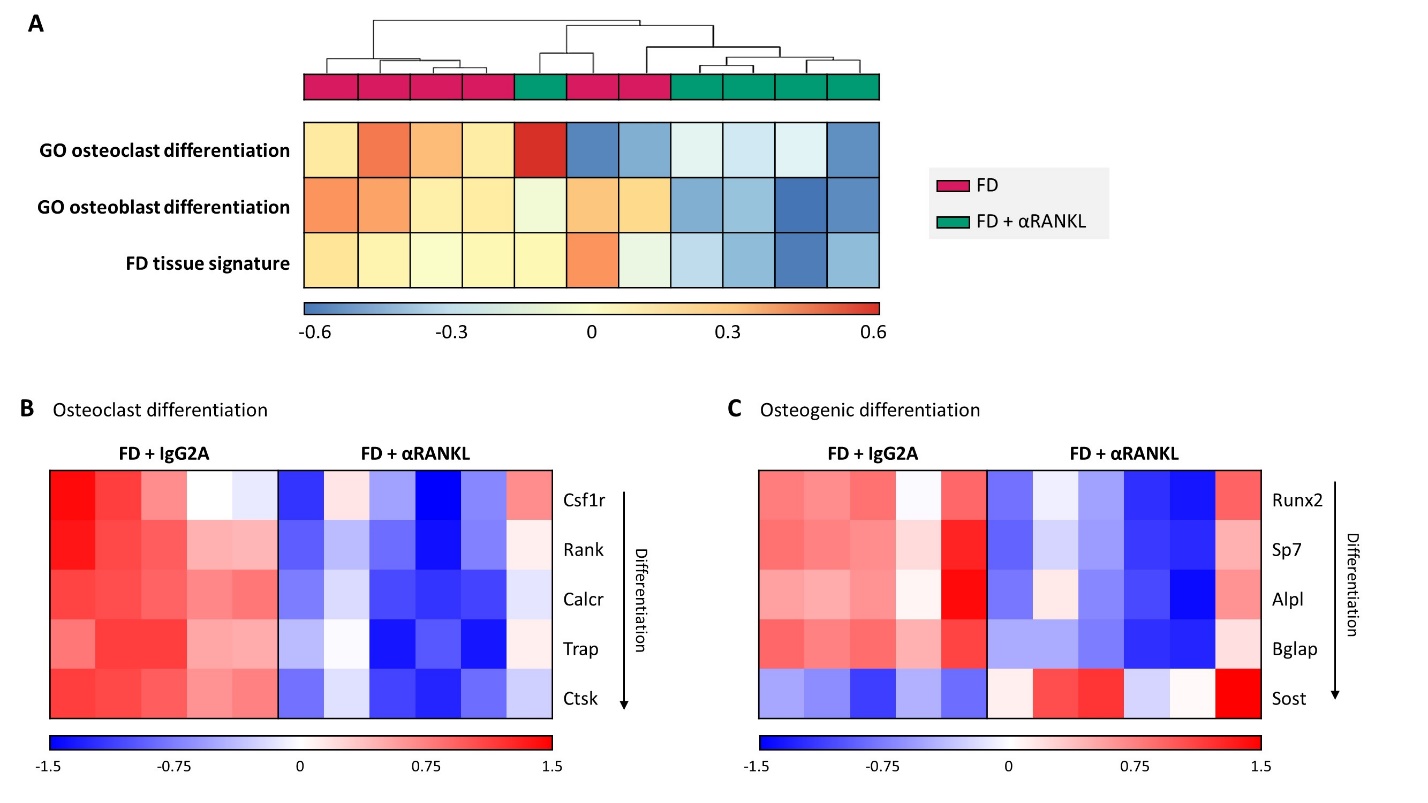

Figure S10 – RNAseq analyses of mouse FD tissue, treated or not with αRANKL. A) Unsupervised clustering of whole transcriptome GSVA scores against selected molecular signatures and genesets. B)

Heatmaps of selected, well characterized gene markers of different stages in osteclastic differentiation and osteoblastic differentiation. All genes related to osteoclastic differentiation where consistently downregulated. Early and late markers of osteoblastic differentiation were significantly downregulated and osteocyte terminal differentiation marker Sost was significantly upregulated after denosumab, consistent with the increased maturation status observed in human FD after denosumab treatment.

#### Figure S11 – *Ex vivo* model of fibrous dysplasia

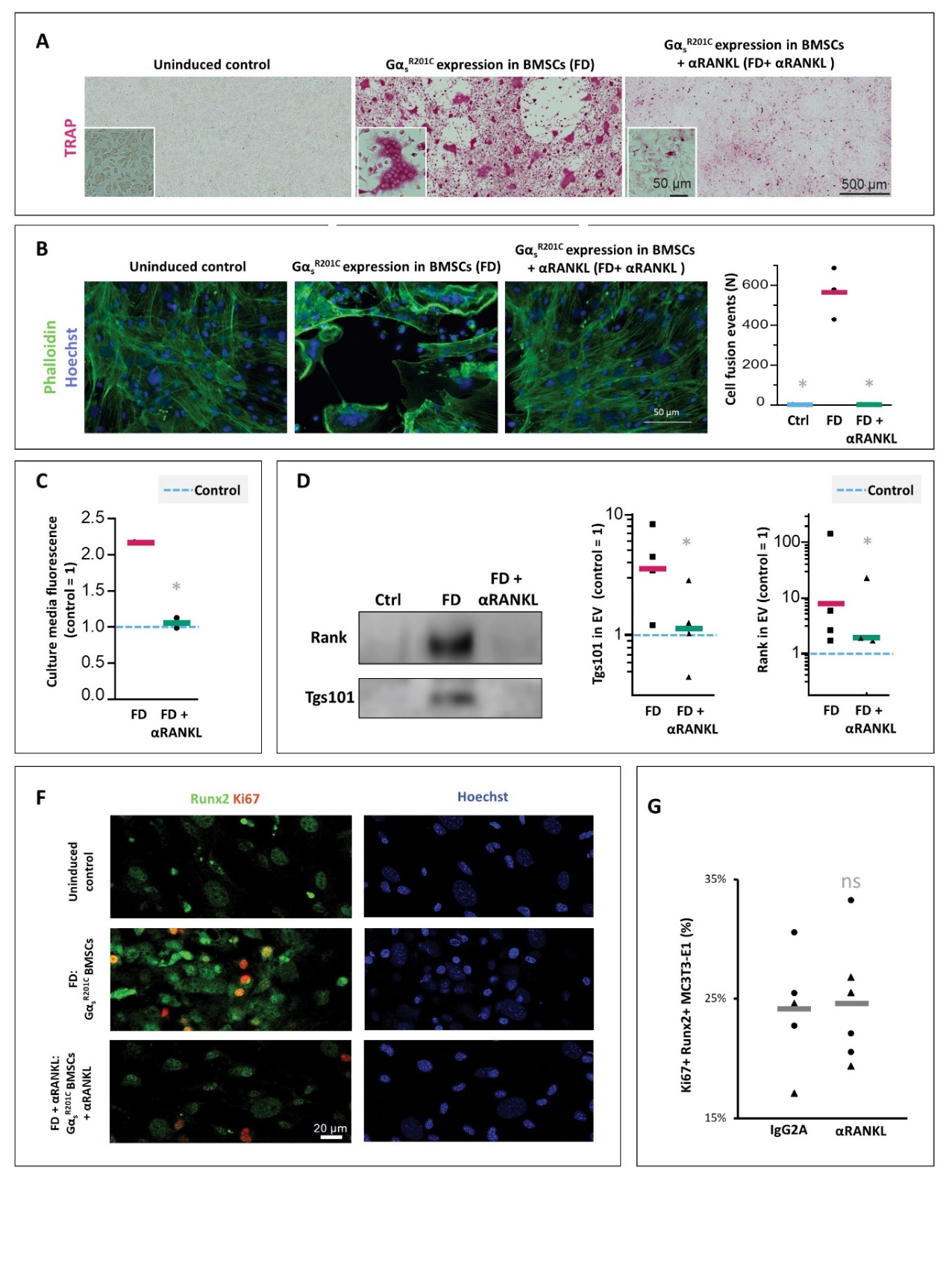

Figure S11 – Ex vivo model of fibrous dysplasia. A) Upon administration of doxycycline in the media, bone marrow cultures develop large TRAP+ osteoclasts and when αRANKL is administered, they dissapear. B) Osteoclast fusion efficiency assay showing no significant cell fusion events in uninduced (Ctrl) and αRANKL-treated cultures. C) In vitro bone resorption assay showing detection of resorption degradation product in the media of induced cultures in comparison to levels in uninduced cultures (blue dashed line). αRANKL administration normalized these levels. D) Western blot and densitometry of extracellular vesicle (EV) lysates obtained from conditioned media of uninduced control (ctrl, blue dashed line) and induced FD cultures with or without αRANKL treatment. RANKL receptor, RANK and EV marker Tgs101 were detected. Lysates from control and treated cultures presented a dramatic decrease in the detection of these markers. F) Nuclear Hoechst stain for the immunofluorescence images shown in Fig. 4E. G) αRANKL administration did not change the abundancy of Ki67+ cells within the Runx2+ cells in cultures of MC3T3-E1 clone 4 (triangles) and 14 (dots) pre-osteoblasts. Data is shown as individual cultures (dots) and group averages (bars). * is p<0.05 vs FD group
